# Implementation of an online tele-coaching community-based exercise (CBE) intervention among adults living with HIV in Canada: A two-phased intervention study

**DOI:** 10.64898/2026.04.02.26350024

**Authors:** Kelly K. O’Brien, Kiera McDuff, Lisa Avery, Francisco Ibáñez-Carrasco, Soo Chan Carusone, Ada Tang, Ahmed M. Bayoumi, George Da Silva, Tai-Te Su, Mona R. Loutfy, Puja Ahluwalia, Darren A. Brown, Patricia Solomon, Ivan Ilic, Zoran Pandovski, Annamaria Furlan, Helen Trent, Mehdi Zobeiry

## Abstract

**Introduction:** Our aim was to examine the implementation of an online community-based exercise (CBE) intervention with adults living with HIV.

**Methods:** We conducted a 12-month community-engaged intervention study with adults living with HIV in partnership with the Toronto YMCA, Canada. We conducted a two phased intervention study involving Phase 1) Intervention: participants were asked to exercise three times/week, supervised every two weeks with online personal coaching, and attend monthly online educational sessions (6-months), and Phase 2) Follow-Up: participants were asked to continue exercising thrice weekly, independently (6-months). We assessed engagement in physical activity (PA) weekly, and body composition, strength, physical function, and flexibility outcomes every two months (bimonthly) across both phases (12-months). We used segmented regression to assess the change in outcomes within and between phases to assess adoption, effect and maintenance of the intervention.

**Results:** Of the 32 participants who initiated, 22 (69%) completed the intervention; and 18 (56%) completed the follow-up. The majority identified as men (69%), median age was 53 years (25th, 75th percentiles: 43, 60), with a median of 3 (1,7) concurrent health conditions. Median number of coaching sessions attended was 10/13 (77%). Participant engagement in ≥30min of moderate or vigorous physical activity in the past week increased from 3.24 days at baseline (95%CI:2.69, 3.79) to 3.36 days (95%CI:2.83,3.89) at the end of intervention to 3.35 days (95%CI:2.81,3.89) at end of follow-up [overall mean increase of 0.11 days (95%CI: 0.02,0.20)]. During the intervention, there were significant mean decreases for weight (-1.31kg), body mass index (BMI) (-0.40kg/m^2^), and waist circumference (-2.92cm); and mean increases for push-ups (+7.11 in a minute), plank time (+38.13 sec), sit-to-stand (+2.86 times in 30 sec), and sit-and-reach (+3.47 cm). There were no changes in muscle mass, body fat percent and fat free mass. During the follow-up phase, there were additional significant mean decreases in body weight (-1.52 kg), BMI (-0.50 kg/m^2^) and sit-to-stand (+1.52 times in 30 sec).

**Conclusions:** Participants demonstrated increases in physical activity and improvements in strength, weight, body composition, physical function, and flexibility with the online CBE intervention, some of which were sustained at the end of the study. Future research should incorporate strategies to enhance engagement in physical activity among adults with HIV.

## Introduction

HIV is a chronic illness, where individuals can experience health-related consequences of HIV, aging and multimorbidity, known as disability [1–3]. Exercise, defined as planned, structured physical activity with the aim to improve or maintain physical fitness is a specific rehabilitation self-management intervention that can address disability among people living with HIV [4]. Performing exercise at least three times per week for at least four weeks can lead to improvements in cardiopulmonary fitness, strength, body composition, and mental health among adults living with HIV [5, 6]. Despite the benefits, engagement in regular exercise varies among adults living with HIV. Evidence from a systematic review reported that 51% of adults living with HIV achieved recommended guidelines of ≥150 minutes of moderate to vigorous aerobic physical activity (any bodily movement resulting in energy expenditure) per week and were less active than others with chronic illness [7].

Community-based exercise (CBE) is a rehabilitation intervention that can improve health among people living with HIV within a self-management framework [8]. Community-based exercise can foster social interaction, peer support, encouragement to exercise, and can promote emotional, cognitive and behavioural self-management strategies to help independently manage chronic health challenges [9, 10]. In earlier work, we evaluated a CBE intervention in which adults living with HIV were expected to engage in thrice weekly exercise for six months, with weekly in-person supervised personal training at the YMCA in Toronto, Canada [11–14]. Few women (<10%) participated in the study [11], citing geographical, childcare, social and interpersonal barriers to exercise in traditional gym settings [13].

Tele-rehabilitation involving the delivery of programs and services via web-based platforms, and specifically online exercise, have emerged as an approach for people living with chronic conditions to engage in physical activity [15]. Online tele-coaching has been well-established for individuals living with neurological conditions [9, 16–18], persistent pain [19], chronic obstructive pulmonary disease [20–22], renal disease [23–27], and inactive adults [28], and offer viable alternatives for engaging in exercise [29]. However, it is unclear how online exercise interventions translate with persons living with HIV.

Our aim was to examine the implementation of an online tele-coaching CBE intervention among adults living with HIV. Using the RE-AIM (Reach-Effectiveness-Adoption-Implementation-Maintenance) Framework [30], specific objectives in this paper were to describe: 1) the extent adults living with HIV participated in the intervention (Reach and Adoption); and 2) impact of the intervention on engagement in physical activity, and health and fitness outcomes over time (Effect & Maintenance).

## Materials and Methods

### Study Design

We used the RE-AIM Framework, to evaluate the implementation of online CBE, and ability of adults living with HIV to integrate exercise into their daily lives over time. We conducted a longitudinal intervention study involving a 6-month online CBE intervention, followed by a 6-month follow-up independent phase. See the previously published protocol for further details [31]. The study protocol was approved by the University of Toronto Research Ethics Board (Protocol # 40410).

Qualitative findings are previously published, reporting on the process of implementation from the perspectives of adults living with HIV and community fitness providers [32]. Goal attainment at the end of the intervention and follow-up phases of the study have been published [33]. In this manuscript we focus on the primary outcome (physical activity) and outcomes related to reach, maintenance and effect of the intervention on physical health outcomes.

### Patient and Public Involvement

This study involved a collaboration with the Toronto Central YMCA, Toronto, Ontario Canada, community-based organizations, a community HIV clinic in collaboration with the Ontario HIV Treatment Network Cohort Study (OCS), and Realize, a national organization focused on advancing education, practice and policy on HIV and rehabilitation [12–14, 34, 35]. Our team includes persons living with HIV, representatives from community-based organizations, clinicians, researchers, and recreation fitness providers who advised on all stages of the research. An Engagement Coordinator living with HIV provided support to participants pertaining to the technology involved with the intervention throughout the study.

### Consent

All participants provided informed verbal consent to participate in the study online via Zoom with the research investigator who signed and dated the consent form confirming verbal consent [36].

### Participants & Recruitment

#### Inclusion Criteria

Adults living with HIV in Toronto who: considered themselves medically safe to exercise as determined by the self-administered Physical Activity Readiness Questionnaire (PAR-Q) [37]; and had access to (i) a smart phone, tablet, laptop, or desktop computer; ii) Wi-Fi or Data Internet Plan; and iii) a web-cam or camera within the device, and iv) space in their home to take part in exercise, and were willing to exercise for one year.

#### Recruitment

We recruited via community-based organizations, an HIV clinic via the Ontario HIV Treatment Network Cohort Study (OCS) (Maple Leaf Medical Clinic), and with participants who agreed to be contacted from a prior CBE study with the YMCA [11, 38] between May 17, 2021 and November 9, 2021. Participants of the OCS responded to a question at the end of the OCS questionnaire asking whether they would be interested in learning more about the CBE tele-coaching study. We intentionally recruited women from our in-person CBE study who were interested but unable to participate due to geographical or childcare reasons [11–13]. We aimed to recruit ≥30% cis- and transgender women to achieve proportional gender and geographical representation in the study [39].

### Protocol Set-Up

Prior to the intervention, fitness instructors took part in a knowledge exchange workshop on topics including HIV, rehabilitation and exercise for people living with HIV, goal setting, research procedures, anti-oppression, and trans inclusion [40]. Participants were provided with exercise equipment to engage in the home-based exercise and fitness assessments, including a wooden step, measuring tape, body weight and composition scale, Therabands®, and a wireless physical activity monitor (Fitbit Inspire 2®) [41]. We piloted the set-up of the technology, including (but not limited to); Zoom software [36], web-cam, YMCA membership, web-based questionnaire software, and Fitbit® App [41], with participants and fitness instructors.

### Online Community-Based Exercise Intervention

The 12-month study involved two phases: 1) Intervention: participants were asked to exercise 3 times/week, supervised every two weeks (biweekly) with online coaching with personal trainers from the YMCA, and attend monthly online educational sessions (6-months), and 2) Follow-Up (Maintenance): participants were asked to continue exercising thrice weekly, independently (6-months).

#### Intervention Phase (6 months)

Participants met a personal trainer to assess their goals and establish an individualized tailored home-based exercise program involving a combination of aerobic, resistance, balance and flexibility training for a total of approximately 60 min, 3X/week for 24 weeks. Specifically, the intervention included the following four components: 1) biweekly (once every 2 weeks) 60 min personal online coaching with a certified trainer from the YMCA (13 sessions) who monitored and progressed exercise intensity; 2) weekly online exercise classes with the YMCA, ∼60 min each led by a trainer at the YMCA; 3) a wireless physical activity monitor (WPAM) to self-monitor steps, distance, calories burned, and active minutes [41, 42]; and 4) monthly online self-management education sessions, focused on topics related to self-management and health, and physical activity living with HIV. We used Zoom as our tele-coaching platform [36]. Participants were able to reschedule their personal coaching sessions for reasons such as illness, injury, and unexpected work commitments or life events, to facilitate access to the total of 13 sessions.

#### Follow-Up (Maintenance) Independent Phase (6 months)

Participants were encouraged to continue with independent exercise three times per week. Participants continued to have access to their YMCA online membership, WPAM, and were encouraged to engage in online YMCA group-based exercise classes (synchronous or asynchronously (recordings)) over Zoom [36].

### Data Collection

#### Reach and Adoption – Level of Engagement in the Intervention

We administered the Information and Communication Technologies (ICT) Development Index (IDI) via online interview at enrollment to assess and discuss digital indicators of access to information and technology (mobile telephone, computer, internet and type of internet), use of technology, and skills with technology (literacy) [43–45]. We assessed the level of engagement in the CBE intervention including attendance to the biweekly online personal coaching sessions, weekly enrollment in the group online exercise classes at the YMCA, weekly WPAM use, and attendance to the monthly online educational sessions.

#### Effect and Maintenance

##### Physical Activity (Primary Outcome)

We assessed engagement in physical activity, weekly. Participants were asked to respond to a brief (2 min) online web-based CBE physical activity questionnaire (CBE-PAQ) weekly to document the nature and extent of activity during both phases of the study (12 months) to assess sustained engagement in exercise.

Participants were asked: i) the number of days in the past week engaged in ≥30 min of moderate or vigorous aerobic physical activity *(Single Physical Activity Item: In the past week, on how many days did you do a total of 30 minutes or more of moderate or vigorous physical activity, which was enough to raise your breathing rate?)* [46] and ii) whether they achieved the weekly recommended Canadian Physical Activity Guidelines for *aerobic activity* (defined as engaging in aerobic activity of moderate to vigorous intensity for at least 150 minutes (2.5 hours) in the past week) and *resistance activity* (defined as engaging in muscle strengthening activities at moderate or greater intensity on at least 2 days in the past week) [47].

To assess engagement in exercise for adults living with HIV over time (maintenance) we measured engagement in *physical activity* and *adherence* to the online CBE intervention over 12 months [47]. We measured physical activity by administering the Rapid Assessment of Physical Activity (RAPA) Questionnaire bimonthly (7 time points) [48]. We measured adherence by documenting attendance to the biweekly fitness sessions in the 6-month intervention (total 13 sessions). The Rapid Assessment of Physical Activity (RAPA) is a 9 item self-reported questionnaire that measures physical activity [48]. The RAPA Aerobic score uses items 1-7 and the score is representative of aerobic exercise engagement, classifying participants as sedentary, under-active, under-active regularly (light activities), under-active regular, or active. Higher RAPA scores indicate greater level and intensity of engagement in physical activity. The RAPA demonstrates construct validity with adults living with HIV [49].

##### Physical Fitness and Health Outcomes (Secondary Outcomes)

We assessed cardiopulmonary fitness, body composition, strength, physical function, and flexibility outcomes bimonthly across the intervention (Month 0, 2, 4, 6) and follow-up phase (Month 8, 10, 12) of the study. Personal trainers at the YMCA conducted the objective assessments bimonthly (month 0, 2, 4, 6, 8, 10, 12) with participants at their home remotely online via Zoom [36]. The fitness assessment included: a) *muscle strength and endurance* (maximum plank duration (seconds), and maximum number of push-ups to failure); b) *physical function* (30-second sit-to-stand test) [50]; c) *cardiopulmonary fitness*; (resting heart rate, heart rate immediately and 1-min after the 3-minute bench step test) [51, 52]; d) *weight and body composition*; (body weight (kg), body mass index (BMI) (kg/m^2^), body fat percent (%), waist and hip circumference (cm), waist-to-hip ratio) [53]; and e) *flexibility* (sit and reach test; cm) [54]. The fitness assessment took approximately 1 hour to complete and YMCA staff documented outcomes using a web-based questionnaire.

We administered a demographic questionnaire at baseline to capture personal attributes, and clinical characteristics. The bimonthly self-reported questionnaire assessments took approximately 45-60 minutes to complete.

Throughout both phases, participants were asked to use a wireless physical activity monitor (WPAM) (Fitbit Inspire 2®) as a motivator for engaging in physical activity and to self-monitor steps, distance, calories burned, and active minutes [41].

Each participant received a 12-month YMCA membership with access to online YMCA group exercise classes, and 6 months of biweekly online personal tele-coaching with a YMCA fitness trainer for their participation in the study. Participants kept the Fitbit Inspire 2®[41] and the exercise equipment (Therabands®, body weight and composition scale, wooden step, and tape measure) as a token of appreciation for participating in the study.

### Analysis

#### Reach and Adoption

We assessed the access to, use of, and skills with technology of those who expressed interest to participate. We described baseline demographic differences between those who completed the study as measured by completing a twelve-month assessment vs those who did not.

##### Engagement in the Intervention

We calculated the proportion of biweekly individual personal coaching sessions attended, and engagement in the monthly online educational sessions in the intervention phase. Adherence was defined as engaging in ≥75% of the 3 weekly exercise sessions throughout [55]. We documented reasons for lack of adherence, and reasons for rescheduling of coaching sessions to capture the episodic nature of health and disability. We also documented enrollment in weekly online group exercise classes at the YMCA and WPAM use throughout the 12-month study.

### Effect and Maintenance

We used segmented regression to assess the change in trend (slope) between the intervention and follow-up phases to assess adoption, effect and maintenance of the intervention [56, 57]. All analyses were intention to treat; assuming that intervention was months 0 to 6 and then follow-up began from months 6 to 12.

Continuous outcomes were analyzed using linear mixed-effects models with a random intercept for participants to account for the repeated-measures structure of the data. We fit separate slopes during the intervention phase and the subsequent follow-up phase. Model-based predictions were obtained at baseline, 6 months, and 12 months from the population-level (fixed-effects) portion of the models. Changes in predicted values between time points (baseline to 6 months, 6 to 12 months, and baseline to 12 months) were estimated using linear contrasts of the fixed effects, with 95% confidence intervals calculated using the t-distribution with residual degrees of freedom. All models were fit using restricted maximum likelihood estimation via the nlme package in R [58].

Binary outcomes (achievement of aerobic and resistance physical activity guidelines, and engagement in physical activity) were analyzed using generalized linear mixed-effects models with a logit link function to account for the repeated-measures structure of the data. As with the linear models, models included a random intercept for participant and separate slopes for each phase (intervention and follow-up). Predicted probabilities of each outcome at baseline, 6 months, and 12 months were computed on the log-odds scale and transformed to the probability scale. Changes in predicted probabilities between time points were estimated using the delta method to obtain appropriate standard errors. Binary models were fit using the lme4 package in R [59].

We conducted a secondary exploratory analysis to estimate sex-specific probabilities over the course of the study. Separate sex-specific models were fit including interaction terms between sex and the time parameters. Modelling details are provided in the statistical supplement and model parameters are presented in the supplemental materials. All statistical analyses were conducted using the R version 4.4.0 [60].

See **Supplemental File 1** for additional (statistical supplement) details for modeling continuous and binary outcomes.

We calculated the time engaged in moderate to vigorous aerobic physical activity each week and the proportion of participants classified as ‘underactive’ versus ‘active’ by the RAPA across all time points [48]. We defined maintenance as the ability to sustain a RAPA classification as ‘active’ post-intervention.

### Sample Size

Our sample size was based on feasibility in this pilot study. Our aim was to recruit 40 participants to have 30 adults living with HIV complete the intervention (≥30% cis- and transgender women). Our prior work suggested this number would enable us to achieve our objectives related to assessing the impact of implementing CBE [12, 13].

## Results

We initiated recruitment in May 2021. We initiated the intervention in October 2021, and continued enrollment and intervention initiation until November 2021, and completed data collection in January 2023. One-hundred individuals indicated they would like to learn more about the study as shown by their response to an item at the end of the Ontario HIV Treatment Network Cohort Study questionnaire, or willingness to be contacted about future research related to HIV and rehabilitation from the previous CBE study. We emailed all who expressed interest, of which 37 responded and met with the coordinator, and were screened for eligibility and inclusion. Of the 37 who were screened, 33 (89%) consented and were enrolled in the study, 32 participants (97%) initiated, and 22 (69%) completed the intervention; and 18 (56%) completed the follow-up maintenance phase of the study (**Fig 1**).

**Fig 1.**
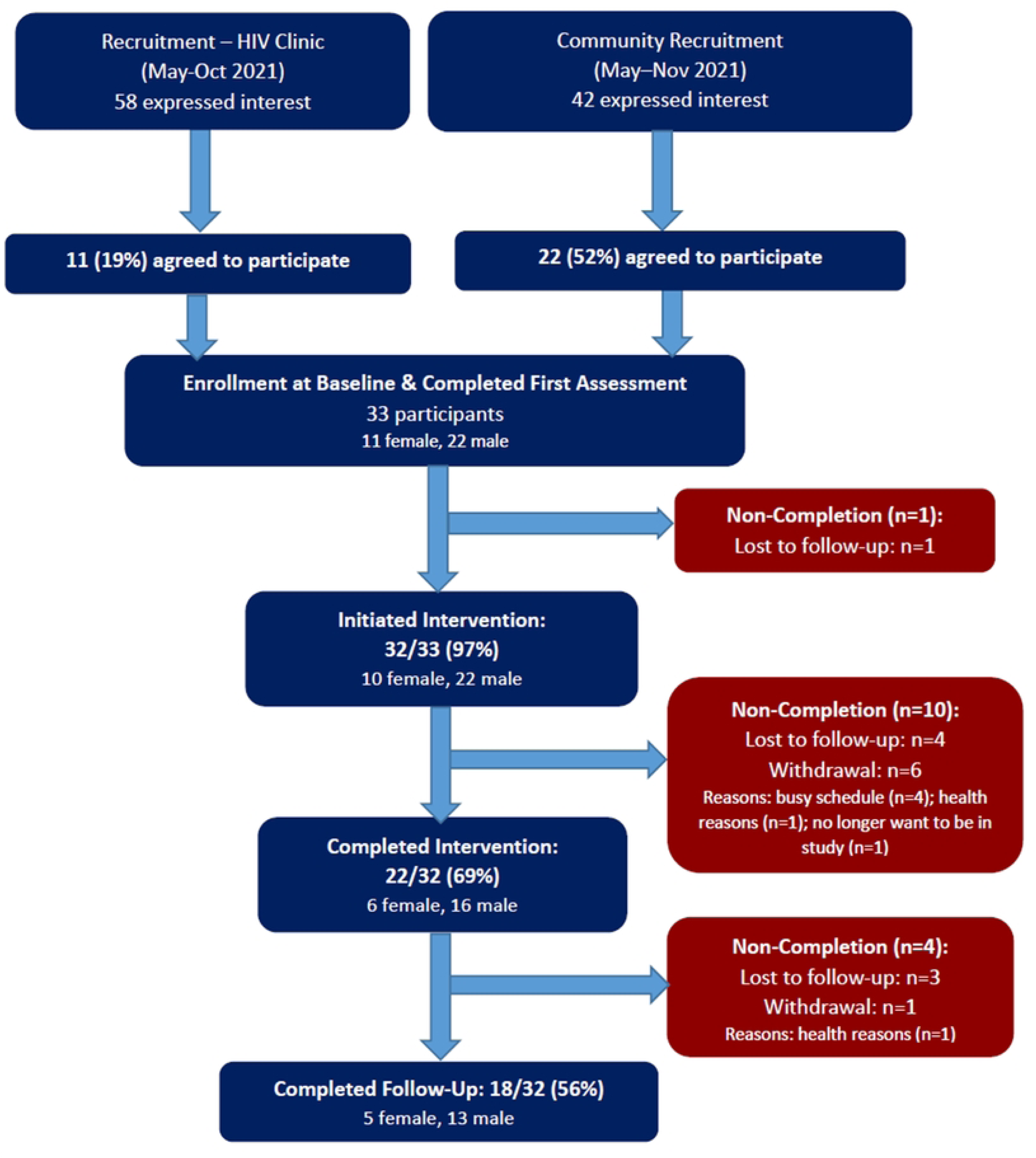
- Participant Flow Chart.

See **Table 1** for characteristics of participants at baseline. The majority were male (69%), median age was 53 years (25th,75th percentiles: 43, 60), with a median of 3 (1, 7) comorbidities in addition to HIV. Nineteen participants (59%) were 50 years or older. Participants were more likely to complete the final 12-month assessment (questionnaire or fitness assessment) if they had completed university, and less likely to complete if they were living with a substance use disorder (concurrent health condition). See **Supplemental file 2** for a comparison of participants who completed versus did not complete a final assessment at study completion (month 12).

**Table 1:**
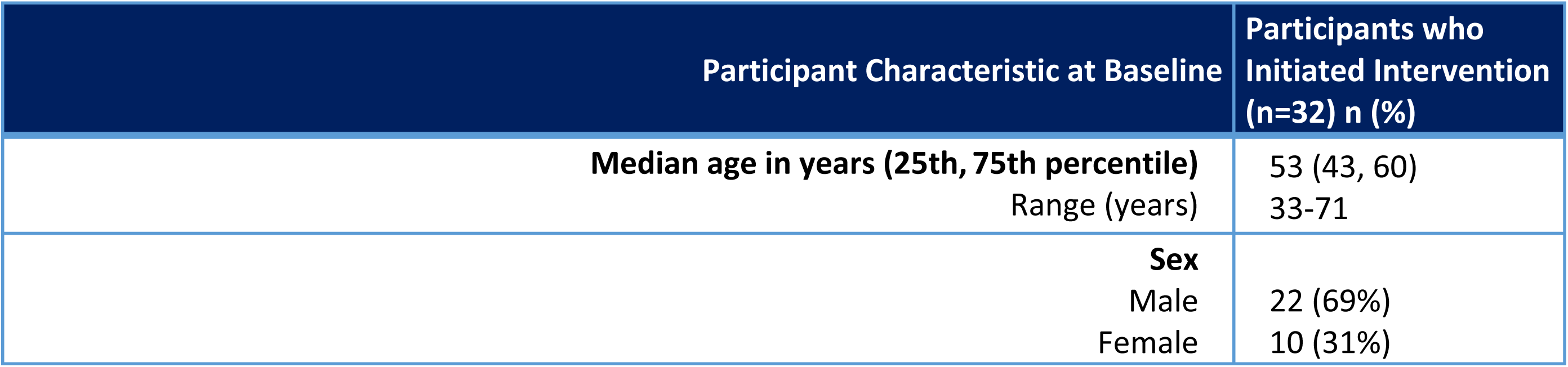
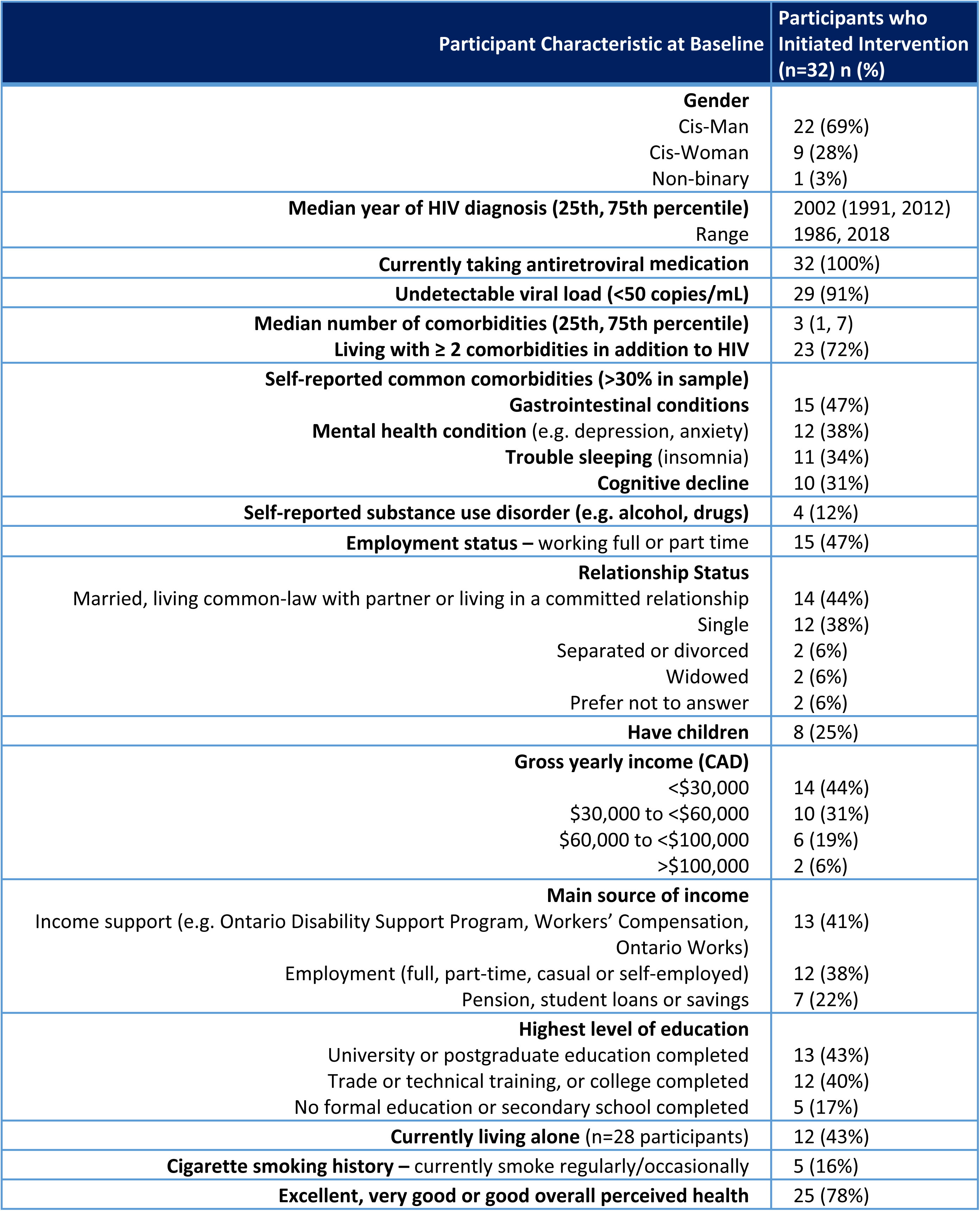
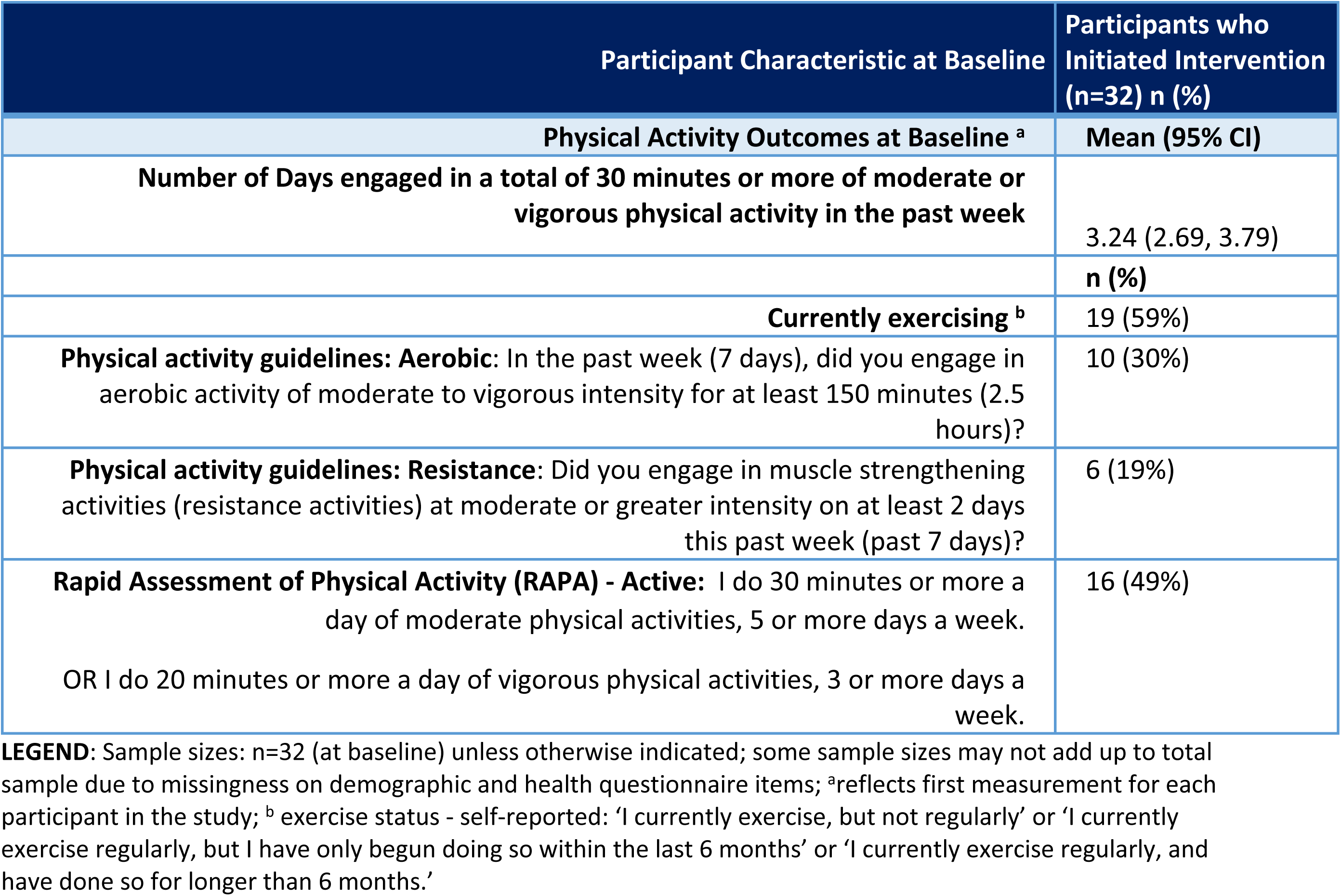
Characteristics of Participants at Baseline (n= 32)

| Participant Characteristic at Baseline | Participants who Initiated Intervention (n=32) n (%) |
| --- | --- |
| Median age in years (25th, 75th percentile)<br>Range (years) | 53 (43, 60)<br>33-71 |
| Sex |  |
| Male | 22 (69%) |
| Female | 10 (31%) |

| **Participant Characteristic at Baseline** | **Participants who Initiated Intervention (n=32) n (%)** |
| --- | --- |
| **Gender** |  |
| Cis-Man | 22 (69%) |
| Cis-Woman | 9 (28%) |
| Non-binary | 1 (3%) |
| **Median year of HIV diagnosis (25th, 75th percentile)** | 2002 (1991, 2012) |
| Range | 1986, 2018 |
| **Currently taking antiretroviral medication** | 32 (100%) |
| **Undetectable viral load (<50 copies/mL)** | 29 (91%) |
| **Median number of comorbidities (25th, 75th percentile)** | 3 (1, 7) |
| **Living with ≥ 2 comorbidities in addition to HIV** | 23 (72%) |
| **Self-reported common comorbidities (>30% in sample)** |  |
| **Gastrointestinal conditions** | 15 (47%) |
| **Mental health condition** (e.g. depression, anxiety) | 12 (38%) |
| **Trouble sleeping** (insomnia) | 11 (34%) |
| **Cognitive decline** | 10 (31%) |
| **Self-reported substance use disorder (e.g. alcohol, drugs)** | 4 (12%) |
| **Employment status – working full or part time** | 15 (47%) |
| **Relationship Status** |  |
| Married, living common-law with partner or living in a committed relationship | 14 (44%) |
| Single | 12 (38%) |
| Separated or divorced | 2 (6%) |
| Widowed | 2 (6%) |
| Prefer not to answer | 2 (6%) |
| **Have children** | 8 (25%) |
| **Gross yearly income (CAD)** |  |
| <\$30,000 | 14 (44%) |
| \$30,000 to <\$60,000 | 10 (31%) |
| \$60,000 to <\$100,000 | 6 (19%) |
| >\$100,000 | 2 (6%) |
| **Main source of income** |  |
| Income support (e.g. Ontario Disability Support Program, Workers' Compensation, Ontario Works) | 13 (41%) |
| Employment (full, part-time, casual or self-employed) | 12 (38%) |
| Pension, student loans or savings | 7 (22%) |
| **Highest level of education** |  |
| University or postgraduate education completed | 13 (43%) |
| Trade or technical training, or college completed | 12 (40%) |
| No formal education or secondary school completed | 5 (17%) |
| **Currently living alone** (n=28 participants) | 12 (43%) |
| **Cigarette smoking history – currently smoke regularly/occasionally** | 5 (16%) |
| **Excellent, very good or good overall perceived health** | 25 (78%) |
| **Physical Activity Outcomes at Baseline <sup>a</sup>** | **Mean (95% CI)** |
| **Number of Days engaged in a total of 30 minutes or more of moderate or vigorous physical activity in the past week** | 3.24 (2.69, 3.79) |
|  | **n (%)** |
| **Currently exercising <sup>b</sup>** | 19 (59%) |
| **Physical activity guidelines: Aerobic:** In the past week (7 days), did you engage in aerobic activity of moderate to vigorous intensity for at least 150 minutes (2.5 hours)? | 10 (30%) |
| **Physical activity guidelines: Resistance:** Did you engage in muscle strengthening activities (resistance activities) at moderate or greater intensity on at least 2 days this past week (past 7 days)? | 6 (19%) |
| **Rapid Assessment of Physical Activity (RAPA) - Active:** I do 30 minutes or more a day of moderate physical activities, 5 or more days a week.<br>OR I do 20 minutes or more a day of vigorous physical activities, 3 or more days a week. | 16 (49%) |

**LEGEND:** Sample sizes: n=32 (at baseline) unless otherwise indicated; some sample sizes may not add up to total sample due to missingness on demographic and health questionnaire items; <sup>a</sup>reflects first measurement for each participant in the study; <sup>b</sup> exercise status - self-reported: 'I currently exercise, but not regularly' or 'I currently exercise regularly, but I have only begun doing so within the last 6 months' or 'I currently exercise regularly, and have done so for longer than 6 months.'

### Access to Technology

Participants who initiated the intervention (n=32) reported similar access to, skills, and use of technology and access to a private space and area to exercise compared with individuals who were screened for eligibility and inclusion (n=37) **(Supplemental file 3).**

### Engagement in the CBE Intervention

See **Supplemental File 4** for an overview of engagement in components of the online CBE intervention.

#### Online Coaching Sessions

Participants attended a median of 10 out of 13 (77%) biweekly online coaching sessions over 25 weeks (25th, 75th percentile: 6, 12). Participants completed their coaching sessions on variable schedules at mutually agreed upon times between the participant and trainer. Ten of 32 participants (31%) extended their biweekly online coaching sessions beyond 6 months. Reasons for not attending and/or extending online coaching sessions beyond 6 months included: scheduling issues (holiday break, trainer-participant scheduling conflicts) (n=6), and/or missed session(s) for unknown reasons (n=4), or for health reasons (n=2). See **Supplemental File 4 (Section A)** for an overview of participant attendance at the biweekly online personal coaching sessions.

#### Weekly online group-based exercise classes

Overall, 13/32 (41%) of participants registered for at least one online group-based exercise class during intervention and follow-up phase. Among these 13 participants, they registered for a median of 2 classes per week (Range: 1 to 13) in the intervention phase and median of 3 classes (Range: 1 to 13) in the follow-up phase. **See Supplemental File 4 (Section B)** for an overview of enrollment in weekly online group exercise classes by week in the study.

#### Weekly wireless physical activity monitor (WPAM) use

We obtained Fitbit data from 18/32 (56%) participants in the study, of which 17/18 (94%) used their WPAM at least once during the study. Among the participants who used the WPAM (56%), they used the device a median of 49/52 weeks (min-max: 0-52 weeks), specifically a median of 26/26 (100%) weeks (min-max: 0-26 weeks) during the intervention phase, and median of 24/26 (92%) weeks (min-max: 0-26 weeks) during the follow-up independent phase. See **Supplemental File 4 (Section C)** for an overview of WPAM usage by week.

#### Monthly online evidence-based self-management education sessions

Attendance to the online educational sessions ranged from 69% to 27%. The most attended session was the first, on the topic of welcome to the study and goal setting, attended by 22 (69%) of participants in the study, followed by dietary strategies for health (47%), sleep health and hygiene (41%), yoga and cognitive health in HIV (27%); HIV and chronic pain; the role for physiotherapy (27%), and coaching yourself beyond the intervention (29%). Slide decks https://hivinmotion.ca/tele-coaching-community-based-exercise-study/ and videos from the presentations are openly available online: https://hivinmotion.ca/learning-2019/. **Supplemental File 4 (Section D).**

### Effect & Maintenance

**Table 3** reports outcomes at baseline, end of six-month intervention, and end of six-month independent follow-up phase; and the *change in outcomes* over each phase of the study. See **Supplemental File 5** for the *rates of change* (slope) in outcomes over the six-month intervention and six-month follow up phases. See **Supplemental 6** (change in outcomes) **and 7** (rates of change) for the results of the secondary exploratory analyses by sex.

**Table 3:**
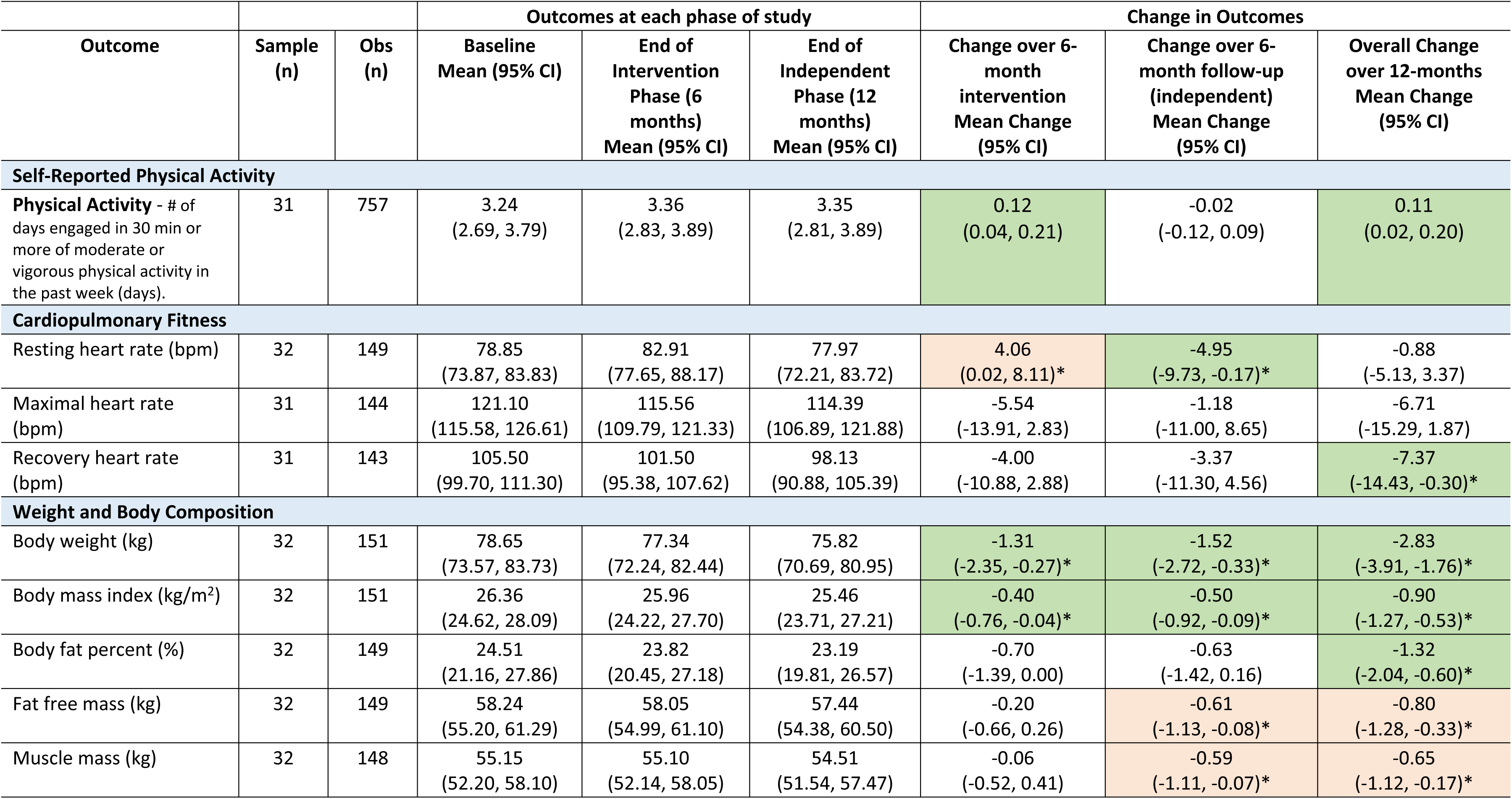
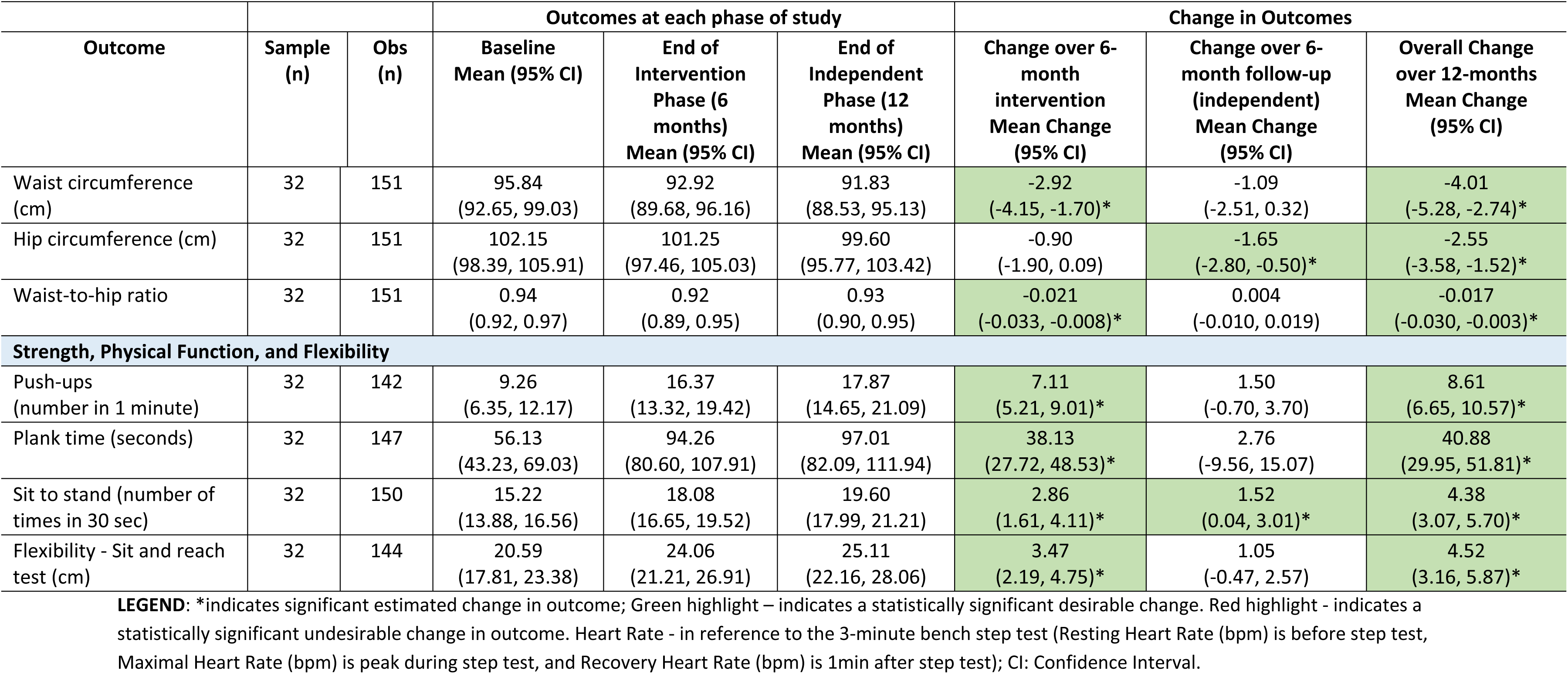
Outcomes estimated at baseline, end of intervention and end of follow-up (independent) phase of the tele-coaching community-based exercise (CBE) Intervention and change in outcomes across each phase for primary and secondary continuous outcomes (n=32 participants)

#### Engagement in Physical Activity – Primary Outcome

##### Number of Days Engaged in Physical Activity in the Past Week

Participant engagement in ≥30min of moderate or vigorous physical activity in the past week significantly increased by 0.12 days per week (95% Confidence Interval (CI): 0.04, 0.21) during the intervention, from 3.24 days at baseline (95%CI: 2.69, 3.79) to 3.36 days (95% CI: 2.83, 3.89) at the end of intervention (week 26), and remaining at 3.35 days (95%CI: 2.81, 3.89) at the end of follow-up (week 52), resulting in an overall significant increase in days engaged in physical activity over the course of the two-phased intervention of 0.11 days per week (95%CI: 0.02, 0.20) (**Table 3, Fig 2**).

**Fig 2.**
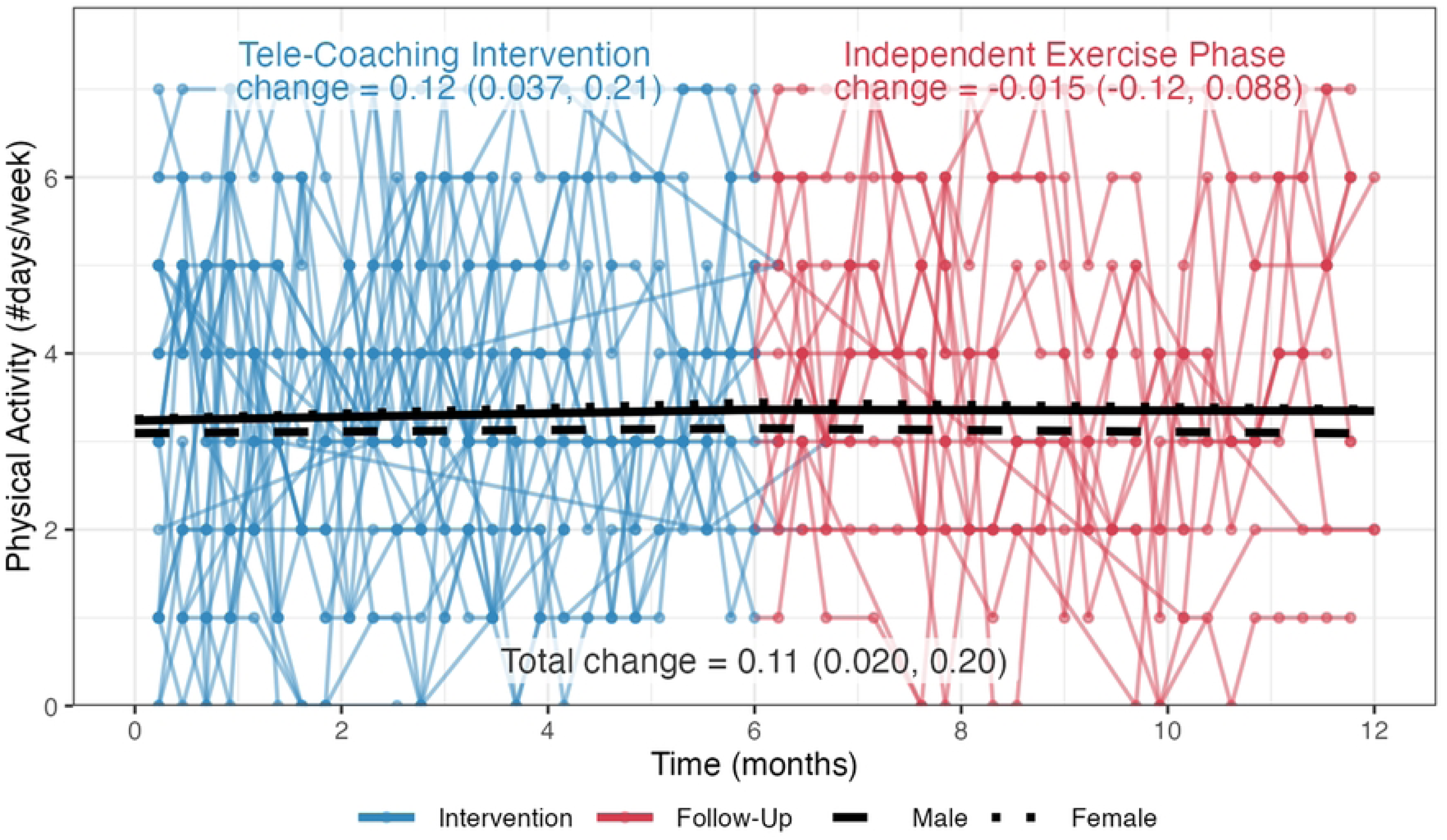
- Number of days engaged in physical activity in the past week across the intervention and independent exercise phases of the study (n=32). *Number of Days of Physical Activity*: “In the past week, on how many days did you do a total of 30 minutes or more of moderate or vigorous physical activity, which was enough to raise your breathing rate? This may include sport, exercise, and brisk walking or cycling for recreation or to get to and from places, but should not include housework or physical activity that may be part of your job.”

#### Achievement of Physical Activity Guidelines

See **Table 4** for estimated probabilities of meeting aerobic and resistance physical activity guidelines and probability of engaging in moderate or vigorous physical activity each week. The probability of achieving physical activity guidelines at baseline was 61% (95%CI: 44%, 75%) for **aerobic activity** (moderate to vigorous intensity for at least 150 minutes (2.5 hours) in the past week) and 65% (95%CI: 49%, 78%) for **resistance activity** (engaging in muscle strengthening activities at moderate or greater intensity on at least 2 days in the past week). The estimated probability of achieving the **aerobic** physical activity guidelines significantly increased by 31% (95%CI: 16%, 46%) over the intervention to 91% (95%CI: 84%, 96%) and significantly reduced by 27% (95%CI: 45%, 9%) during follow-up to 65% (95%CI: 44%, 81%) at the end of the follow-up independent phase. The estimated probability of achieving **resistance** physical activity guidelines did not significantly change over the intervention resulting in 74% (95%CI: 60%, 84%) at the end of the intervention, and significantly reduced by 28% (95%CI: 46%, 11%) during the follow-up phase to 46% (95%CI: 28%, 64%) at the end of the follow-up independent phase of the study (**Table 4**).

**Table 4:** Outcomes estimated at baseline, end of intervention and end of follow-up phase of the tele-coaching community-based exercise (CBE) intervention and change in outcomes across each phase for binary outcomes (n=32 participants)

|  |  |  | Outcomes at each phase of study |  |  | Change in Outcomes |  |  |
| --- | --- | --- | --- | --- | --- | --- | --- | --- |
| Outcome | Sample (n) | Obs (n) | Baseline Probabilities Mean (95% CI) | End of Intervention Phase (6 months) Probabilities Mean (95% CI) | End of Independent Phase (12 months) Probabilities Mean (95% CI) | Change over 6-month intervention Probabilities Mean Change (95% CI) | Change over 6-month follow-up (independent) Probabilities Mean Change (95% CI) | Overall Change over 12-months Probabilities Mean Change (95% CI) |
| Self-Reported Physical Activity |  |  |  |  |  |  |  |  |
| Achievement of physical activity guidelines in the past week: **Aerobic** | 31 | 757 | 0.61 (0.44, 0.75) | 0.91 (0.84, 0.96) | 0.65 (0.44, 0.81) | 0.31 (0.16, 0.46) | -0.27 (-0.45, -0.09) | 0.04 (-0.13, 0.21) |
| Achievement of physical activity guidelines: **Resistance** | 31 | 757 | 0.65 (0.49, 0.78) | 0.74 (0.60, 0.84) | 0.46 (0.28, 0.64) | 0.09 (-0.05, 0.23) | -0.28 (-0.46, -0.11) | -0.19 (-0.35, -0.04) |
| Report of Active as measured by the RAPA scale | 32 | 157 | 0.61 (0.39, 0.79) | 0.79 (0.58, 0.91) | 0.82 (0.57, 0.94) | 0.19 (-0.06, 0.43) | 0.03 (-0.19, 0.25) | 0.22 (-0.02, 0.45) |

**LEGEND: Achievement of Physical Activity Guidelines (Aerobic)** - In the past week (7 days), did you engage in aerobic activity of moderate to vigorous intensity for at least 150 minutes (2.5 hours)? (measured by weekly questionnaire); **Achievement of Physical Activity Guidelines (Resistance)** - Did you engage in muscle strengthening activities (resistance activities) at moderate or greater intensity on at least 2 days this past week (past 7 days? (measured by weekly questionnaire); **Rapid Assessment of Physical Activity (RAPA) Scale (Score of 6 or 7):** 'I do 30 minutes or more a day of moderate physical activities, 5 or more days a week (Score 6)', OR 'I do 20 minutes or more a day of vigorous physical activities, 3 or more days a week (Score 7).' (measured bimonthly). Green highlight – indicates a statistically significant desirable change in outcome. Red highlight - indicates a statistically significant undesirable change in outcome.

#### Cardiopulmonary Fitness

Results for cardiopulmonary fitness outcomes over the intervention and follow-up phases are presented in **Fig 3 to 5** and **Table 3**. There was a significant increase in mean **resting heart rate** at the end of the intervention phase of +4.06 bpm (95%CI: 0.02, 8.11) followed by a significant decrease at the end of the follow-up phase (-4.95 bpm; 95%CI: -9.73, -0.17) resulting in no change across both phases of the study (-0.88 bpm; 95%CI: -5.13, 3.37) (**Fig 3**; **Table 3**).

**Fig 3.**
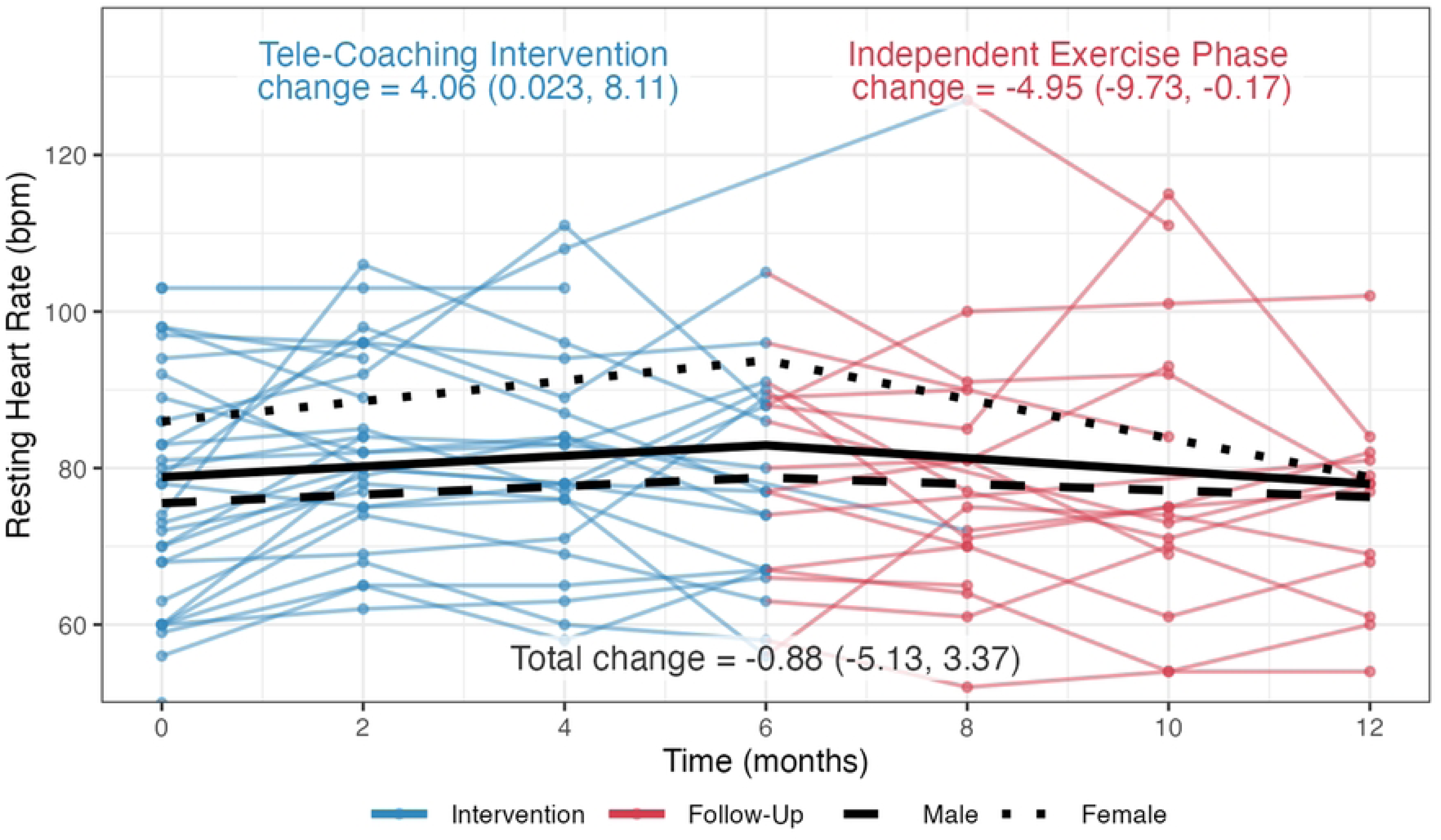
– Cardiopulmonary Fitness Outcome – Resting Heart Rate (beats per minute)

**Fig 4.**
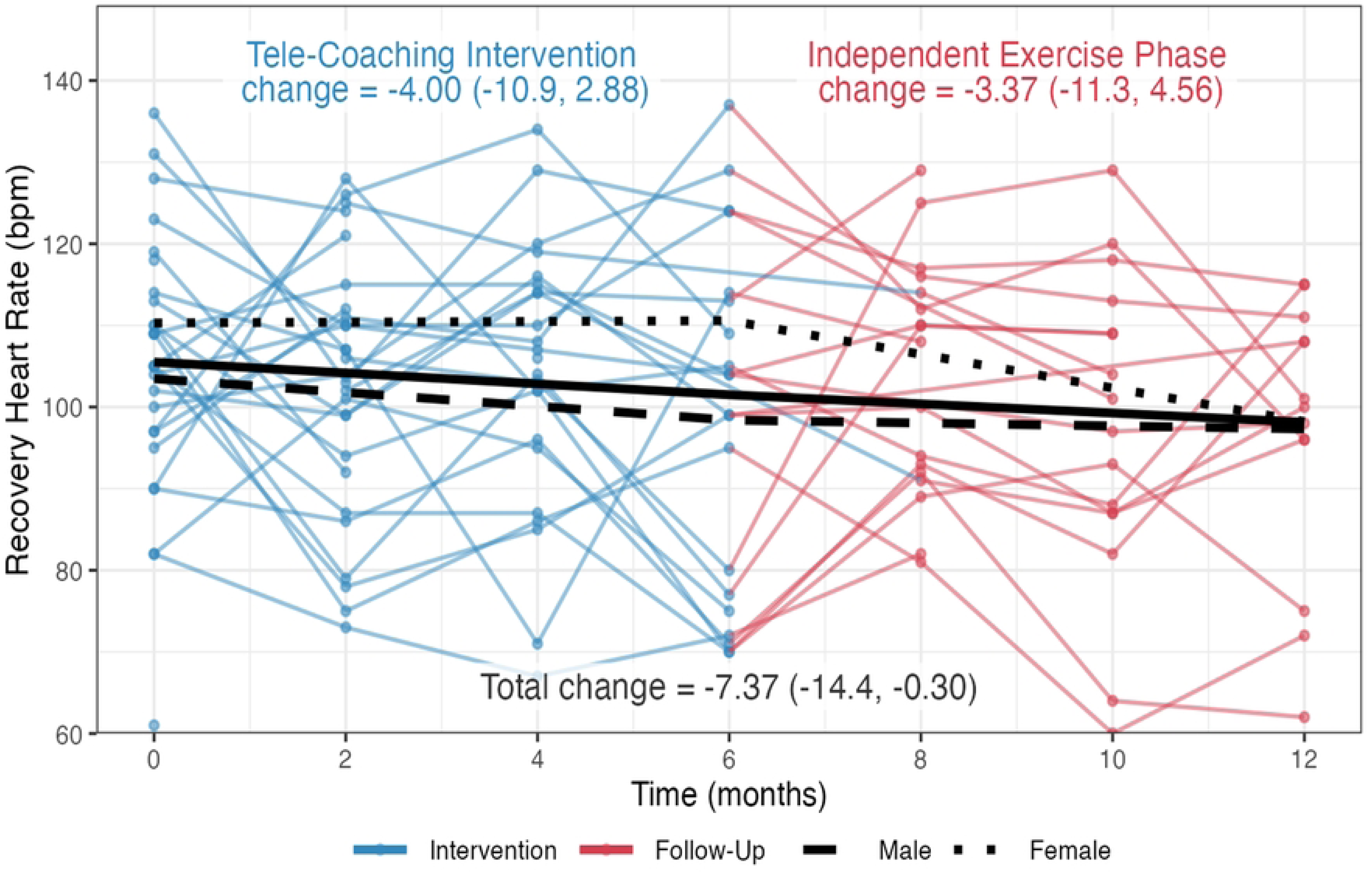
- Cardiopulmonary Fitness Outcome – Recovery Heart Rate (beats per minute)

**Fig 5.**
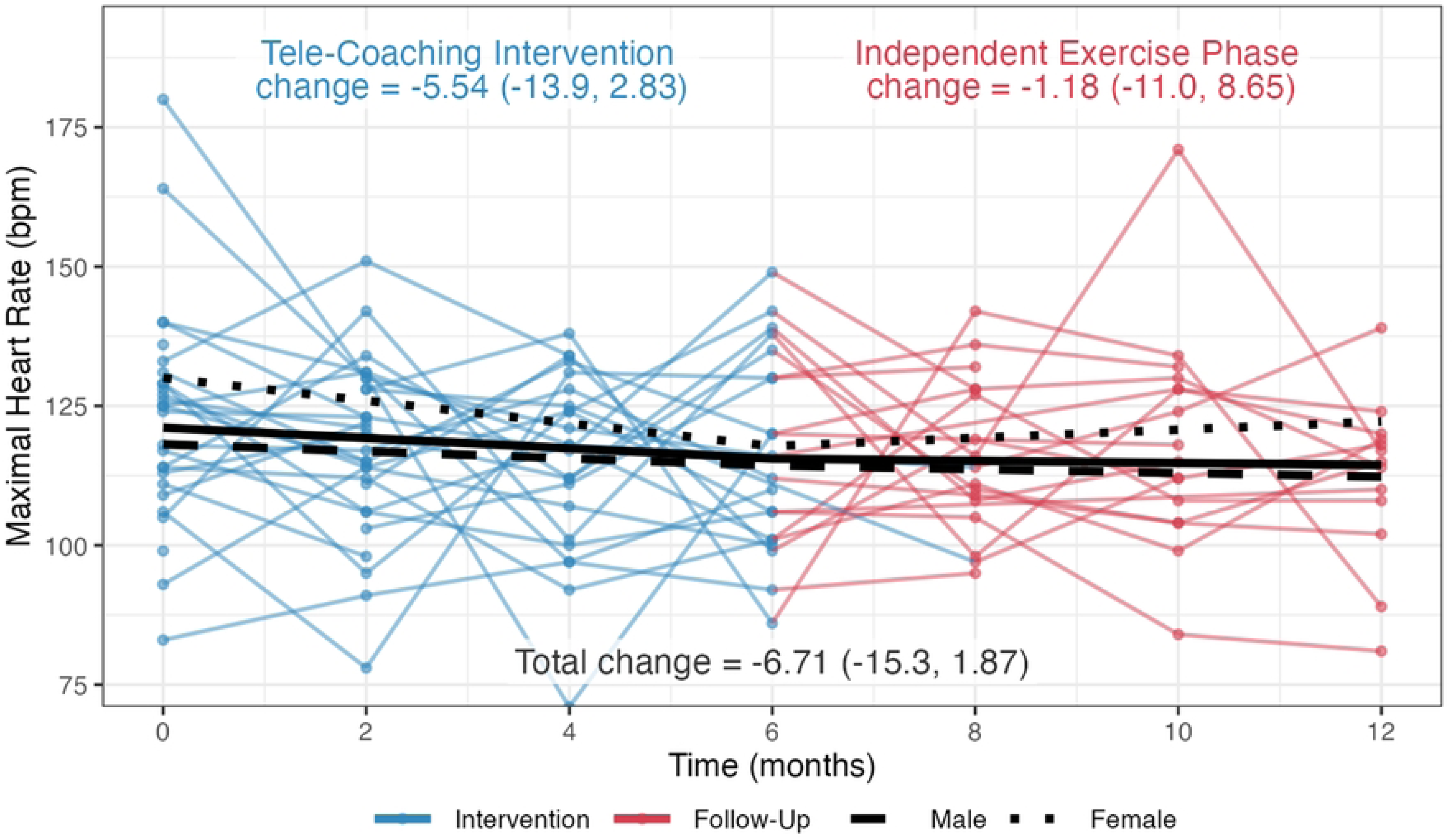
- Cardiopulmonary Fitness Outcome – Maximal Heart Rate (beats per minute)

There was a trend towards a decrease (improvement) in mean **recovery heart rate** at the end of the intervention phase of (-4.00 bpm; 95%CI: -10.88, 2.88) and end of follow-up phase (-3.37 bpm; 95%CI: -11.30, 4.56) resulting in an overall significant mean reduction in recovery heart rate (-7.37 bpm; 95%CI: -14.43, -0.30) across both phases of the study (**Fig 4**; **Table 3**).

Despite a trend towards a decrease (improvement) in mean **maximal heart rate,** there was no significant change over the intervention (-5.54 bpm; 95%CI: -13.91, 2.83), follow-up (-1.18 bpm; 95%CI: -11.00, 8.65) or across both phases (12 months) of the study (-6.71 bpm; 95%CI: -15.29, 1.87) (**Fig 5**, **Table 3**).

#### Weight and Body Composition

Results for weight and body composition outcomes over the intervention and follow-up phases are presented in **Fig 6 to 13 and Table 3**.

**Figure 6.**
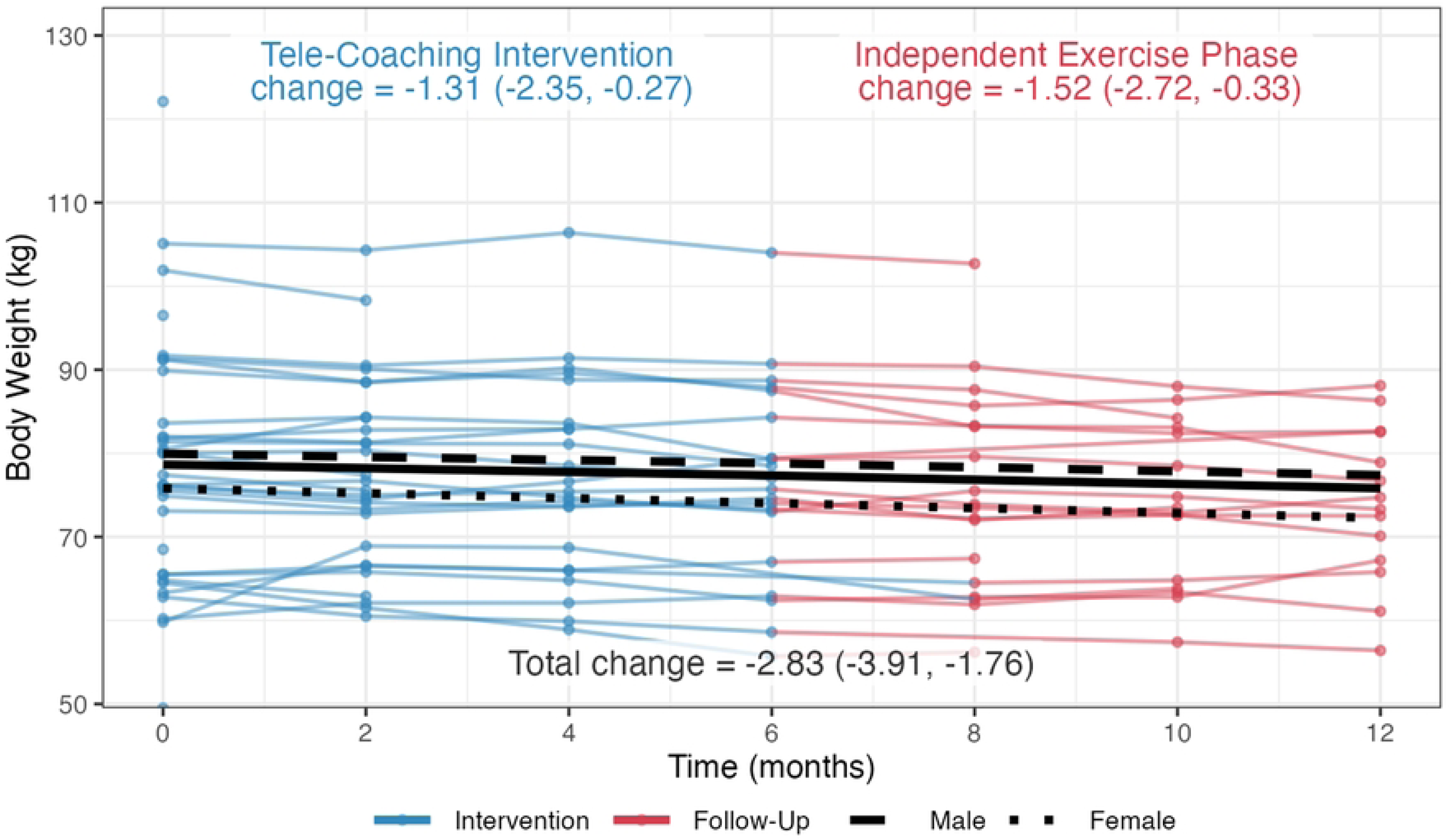
– Weight and Body Composition Outcome – Body Weight (kg)

**Figure 7.**
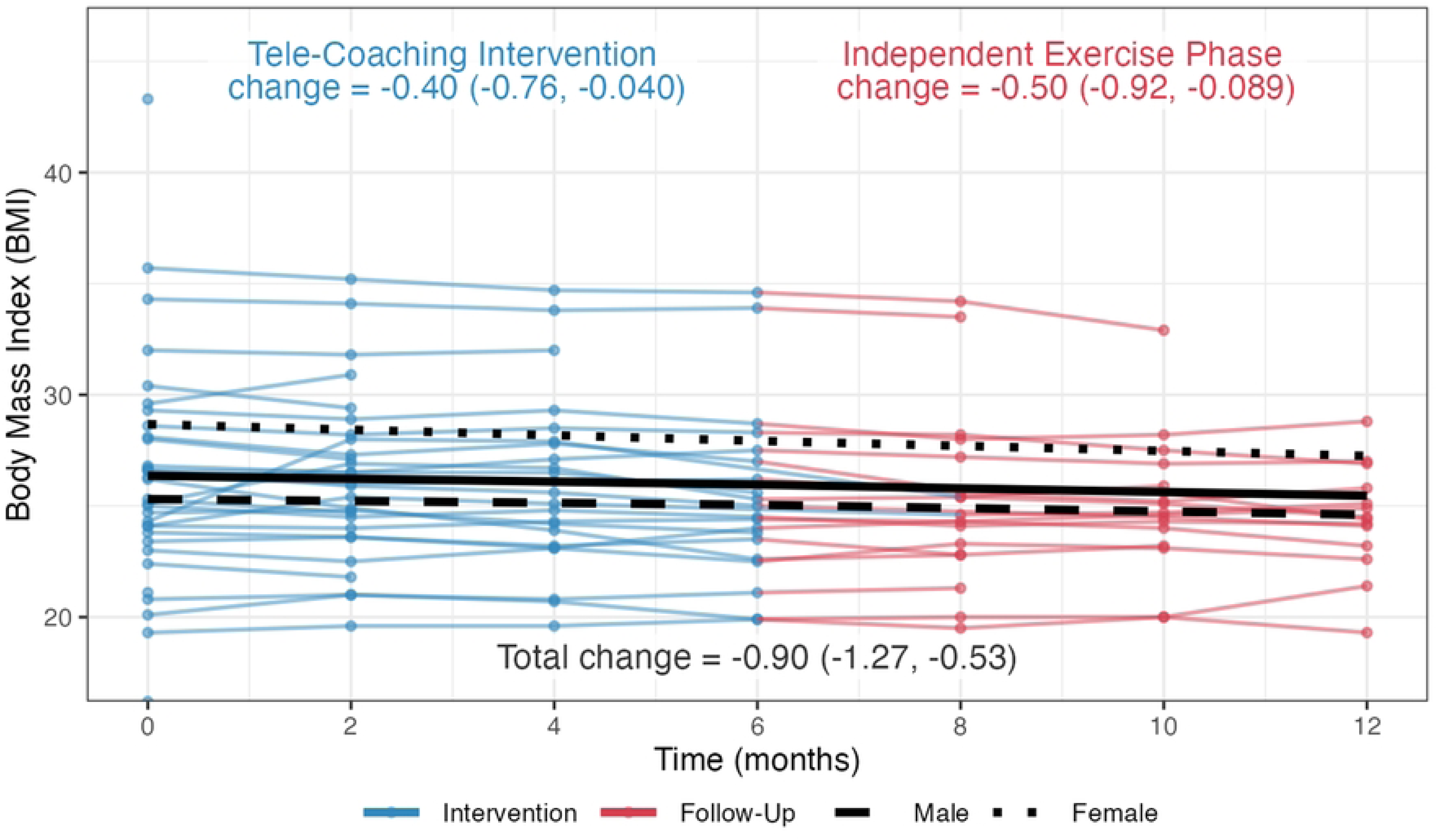
– Weight and Body Composition Outcome – Body Mass Index (kg/m^2^)

**Figure 8.**
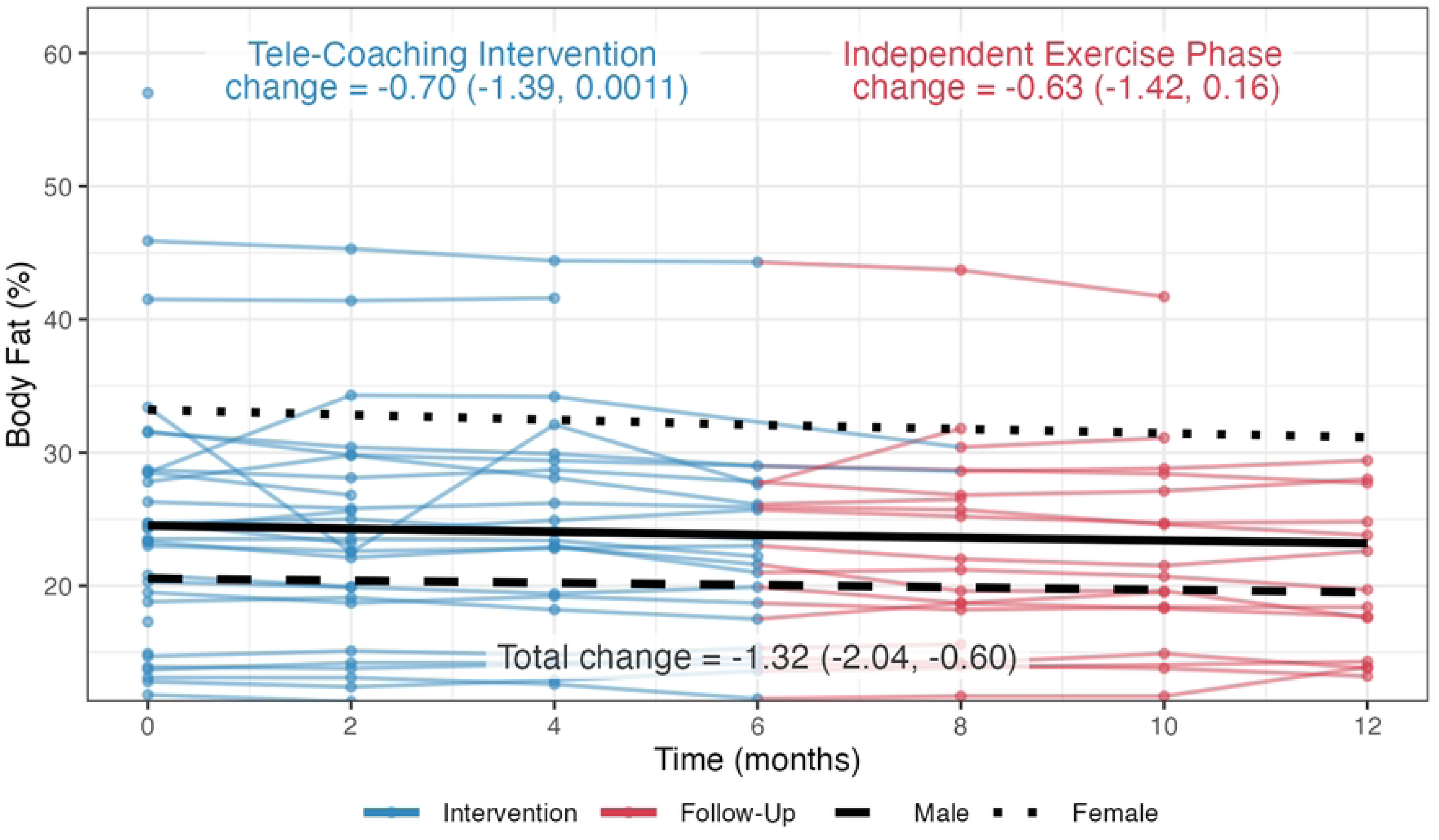
– Weight and Body Composition Outcome – Body Fat Percent (%)

**Figure 9.**
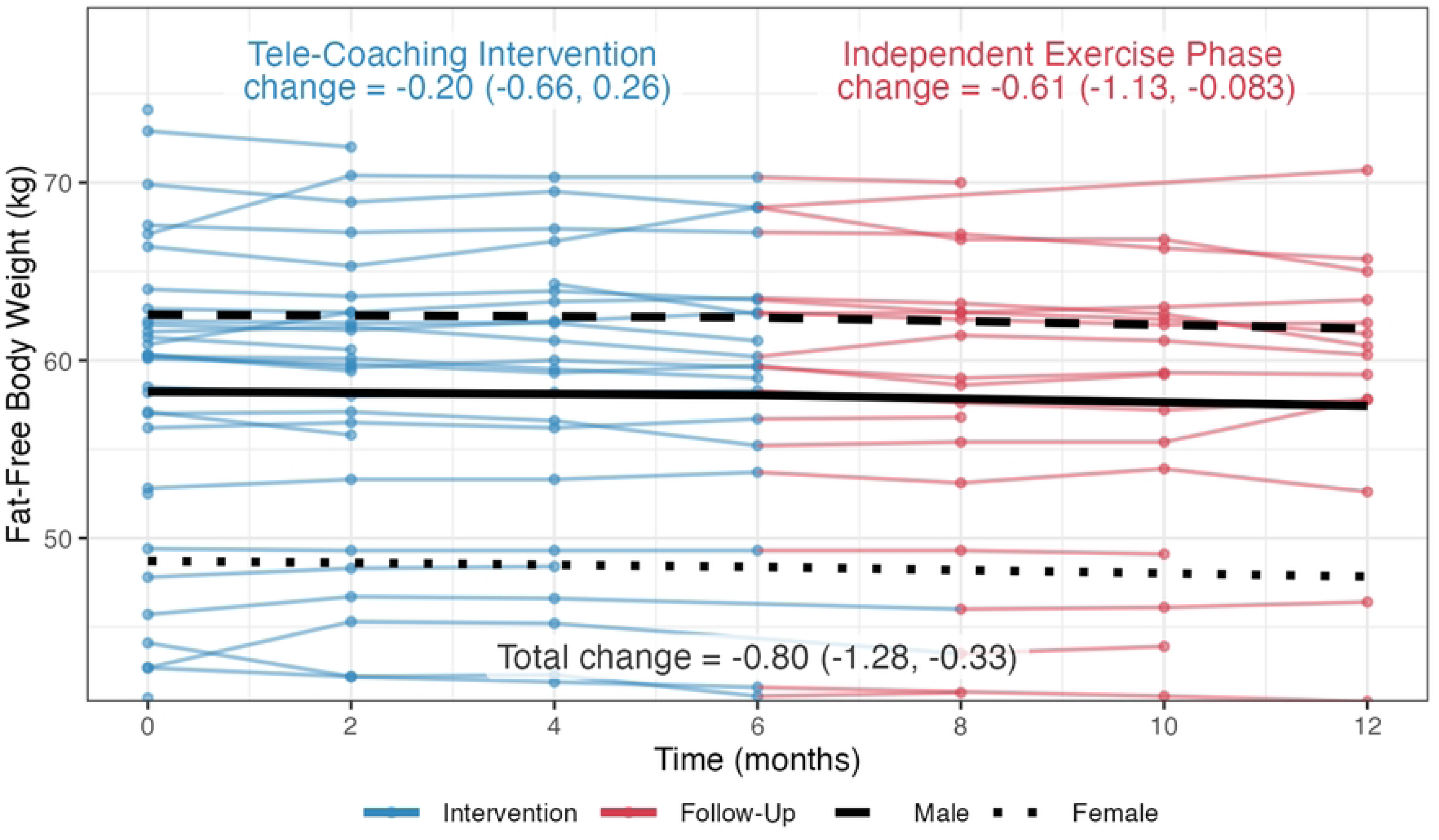
– Weight and Body Composition Outcome – Fat Free Mass (kg)

**Figure 10.**
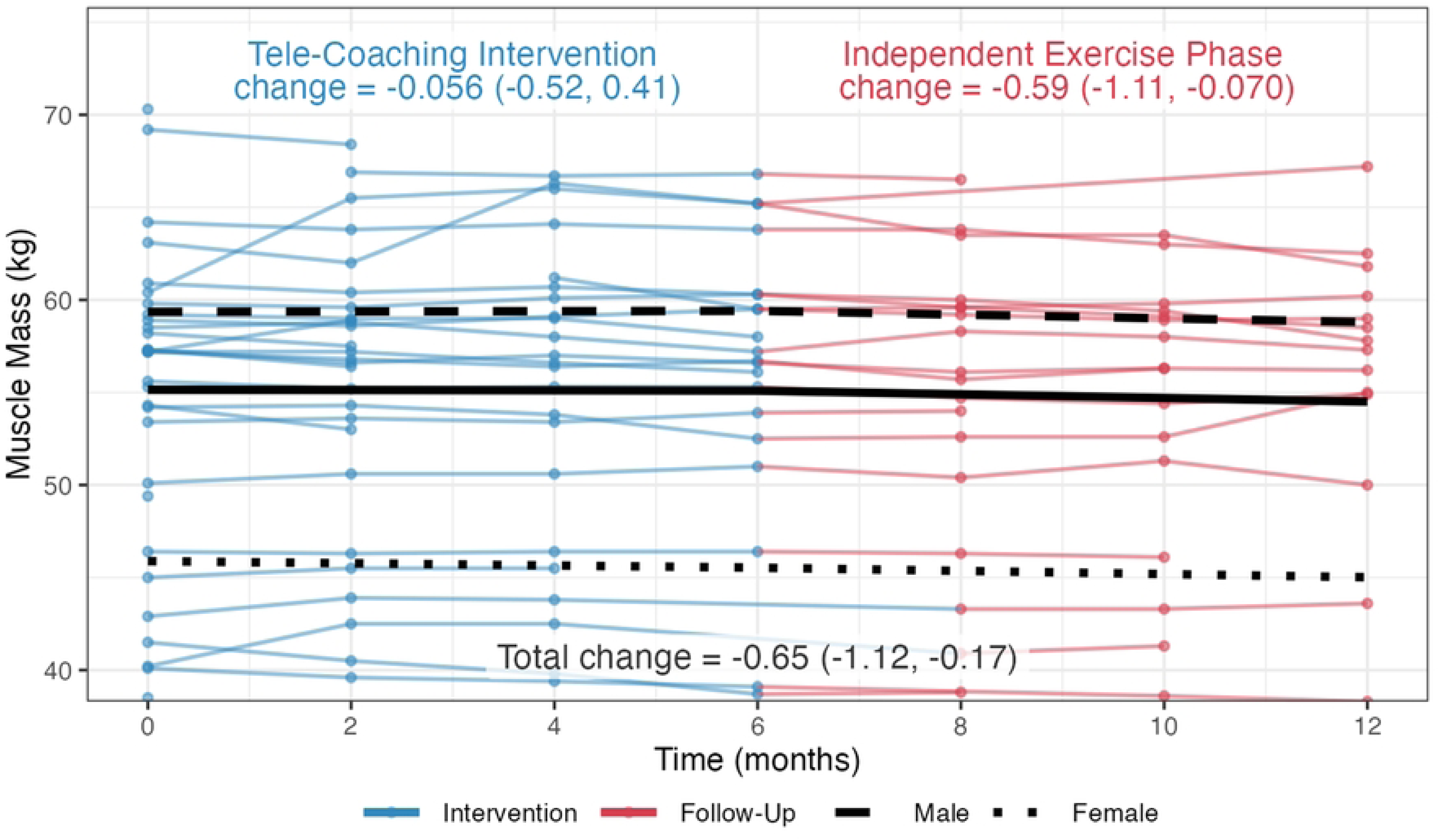
– Weight and Body Composition Outcome – Muscle Mass (kg)

**Figure 11.**
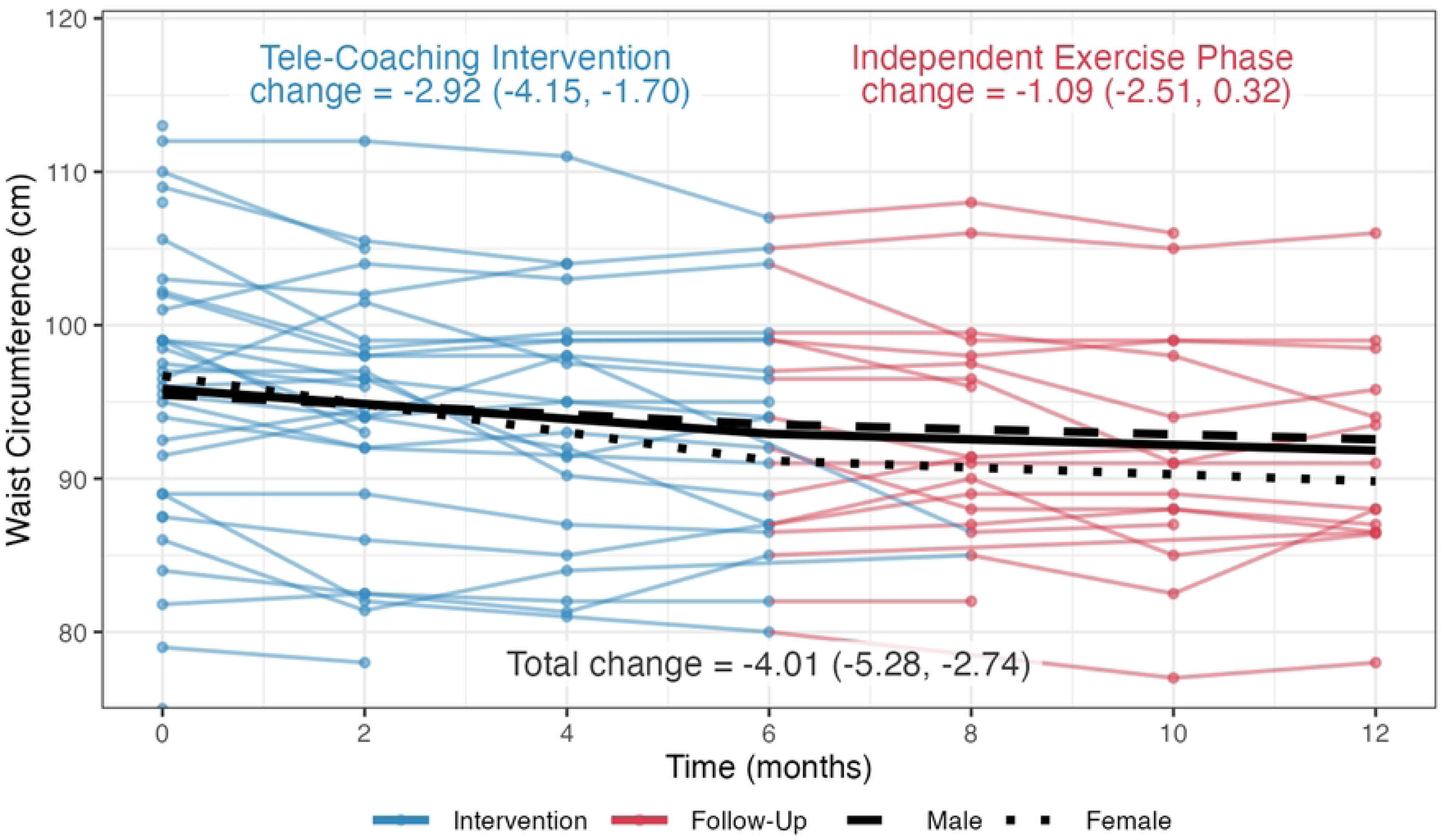
- Weight and Body Composition Outcome – Waist Circumference (cm)

**Figure 12.**
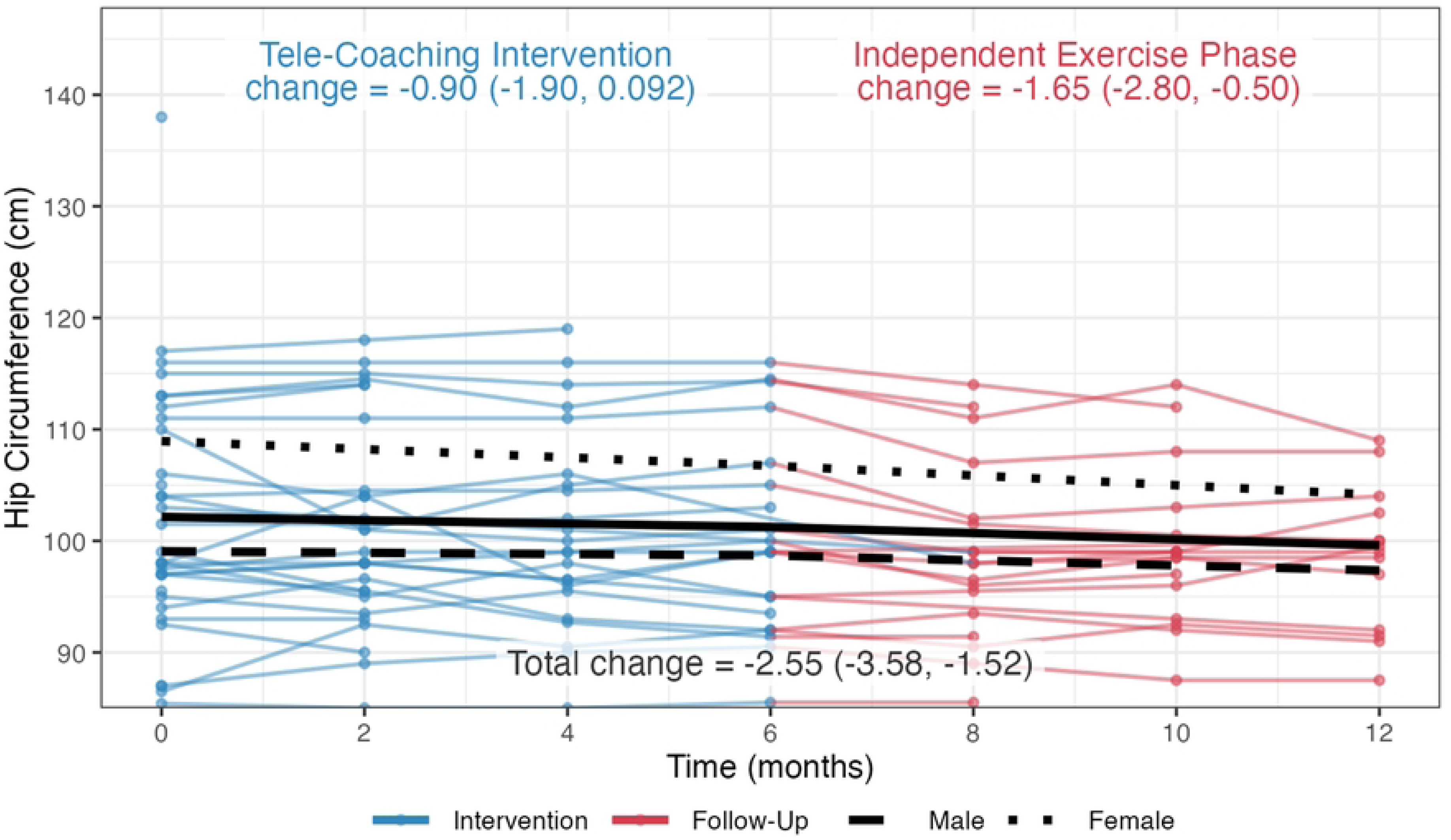
– Weight and Body Composition Outcome – Hip Circumference (cm)

**Figure 13.**
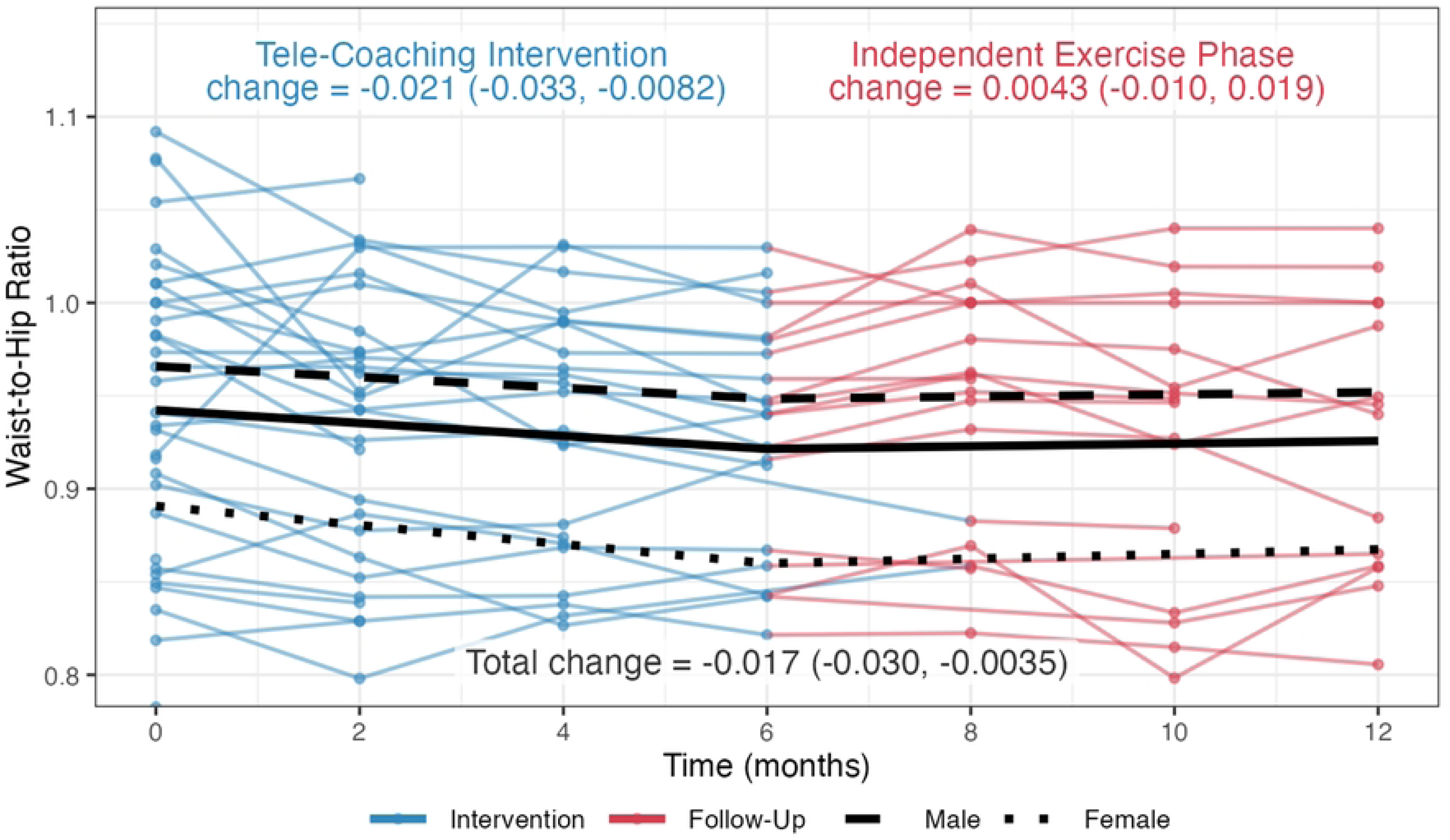
– Weight and Body Composition Outcome – Waist-to-Hip Ratio.

At the end of the 12-month study there were significant mean decreases in all outcomes of weight and body composition. Specifically, at end of the intervention phase, there were significant mean decreases in weight (-1.31kg; 95%CI: -2.35, -0.27), body mass index (BMI) (-0.40kg/m^2^; 95%CI: -0.76, -0.04), waist circumference (-2.92cm; 95%CI: -4.15, -1.70), and waist-to-hip ratio (-0.021; 95%CI: -0.033, 0.008) with significant and sustained decreases for weight (-1.52kg, 95%CI: -2.72, -0.33) and BMI (-0.50kg/m2, 95%CI: -0.92, -0.09) at the end of the follow-up phase (**Table 3**). There were no changes in body fat percent, fat free mass, muscle mass, or hip circumference during the intervention phase, but there were significant reductions in fat free mass (-0.61kg; 95%CI: -1.13, -0.08), muscle mass (-0.59kg; 95%CI: -1.11, -0.07) and hip circumference (-1.65cm; 95%CI: -2.80, -0.50) at the end of the follow-up phase of the study (**Table 3**). Overall changes in weight and body composition across both phases of the study included mean decreases in body weight (-2.83 kg; 95%CI: -3.91, -1.76) (**Fig 6**), BMI (-0.90 kg/m^2^; 95%CI: -1.27, -0.53) (**Fig 7**), body fat percent (-1.32%; 95%CI: -2.04, -0.60) (**Fig 8**), fat free mass (-0.80 kg; 95%CI: -1.28, -0.33) (**Fig 9**), muscle mass (-0.65kg; 95%CI: -1.12, -0.17) (**Fig 10**), waist circumference (-4.01 cm; 95%CI: -5.28, -2.74) (**Fig 11**), hip circumference (-2.55 cm; 95%CI: -3.58, -1.52) (**Fig 12**), and waist-to-hip ratio (-0.017; 95%CI: -0.030, -0.003) (**Fig 13**).

#### Strength, Physical Function & Flexibility

At the end of the intervention phase (6 months), there were significant improvements for all outcomes of strength, physical function, and flexibility. Participants demonstrated mean increases in **push-ups** (+7.11 more pushups in a minute; 95%CI: 5.21, 9.01) (**Fig 14**), **plank time** (+38.13 sec; 95%CI: 27.72, 48.53) (**Fig 15**), **sit-to-stand** (+2.86 times in 30 seconds; 95%CI: 1.61, 4.11) (**Fig 16**), and **sit-and-reach** (+3.47 cm; 95%CI: 2.19, 4,75) (**Fig 17**). (**Table 3**). These improvements were sustained or further increased in the follow up phase (sit to stand) to result in an overall significant increase (improvement) in all outcomes across both phases of the 12-month study for **push-ups** (+8.61 more pushups in a minute; 95%CI: 6.65, 10.57) (**Fig 14**), **plank time** (+40.88 sec; 95%CI: 29.95, 51.81) (**Fig 15**), **sit-to-stand** (+4.38 times in 30 seconds; 95%CI: 3.07, 5.70) (**Fig 16**), and **sit-and-reach** (+4.52 cm; 95%CI: 3.16, 5.87) (**Fig 17**) (**Table 3**).

**Figure 14.**
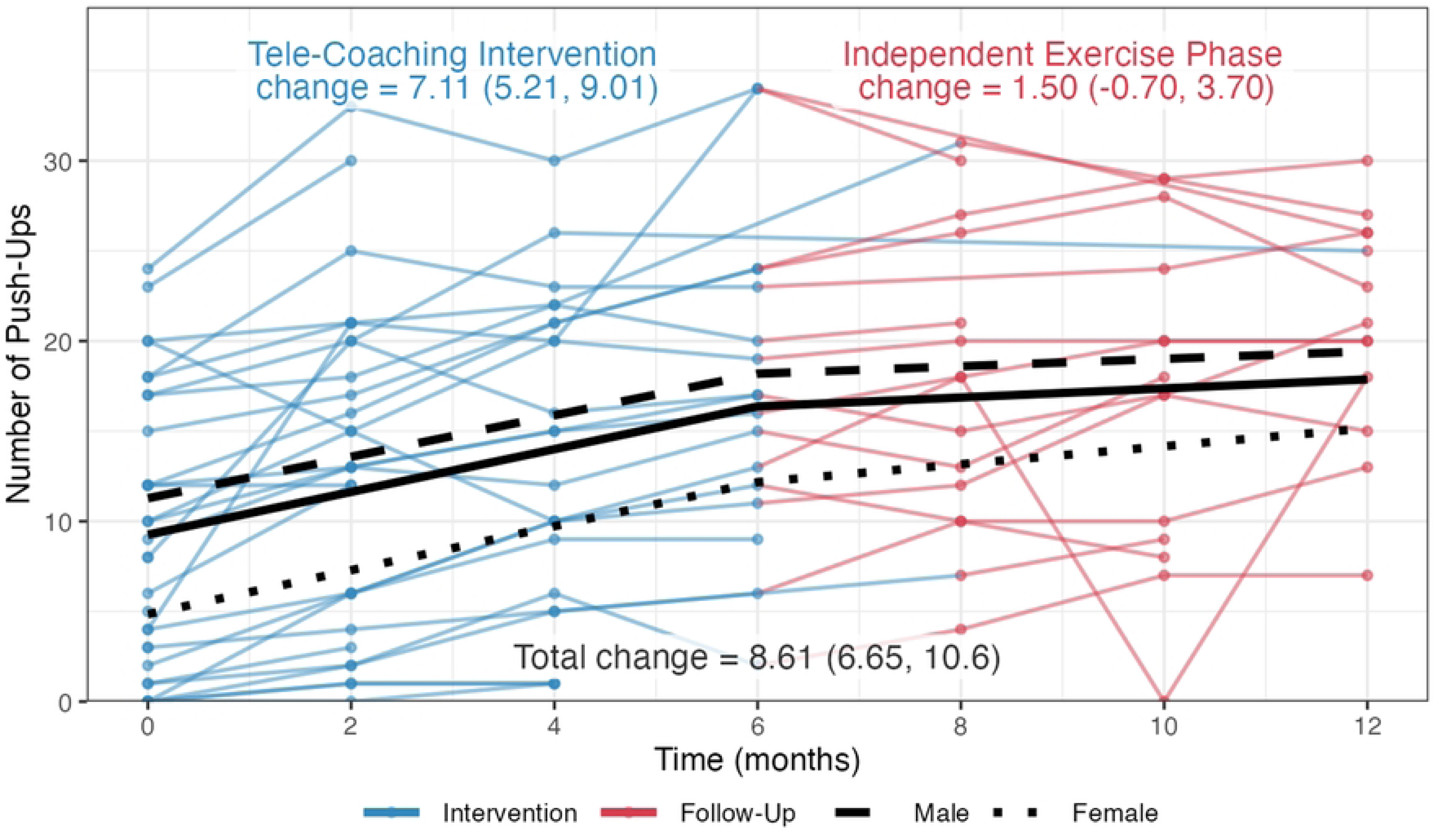
– Strength Outcome – Push-Ups (Number in 1 minute)

**Figure 15.**
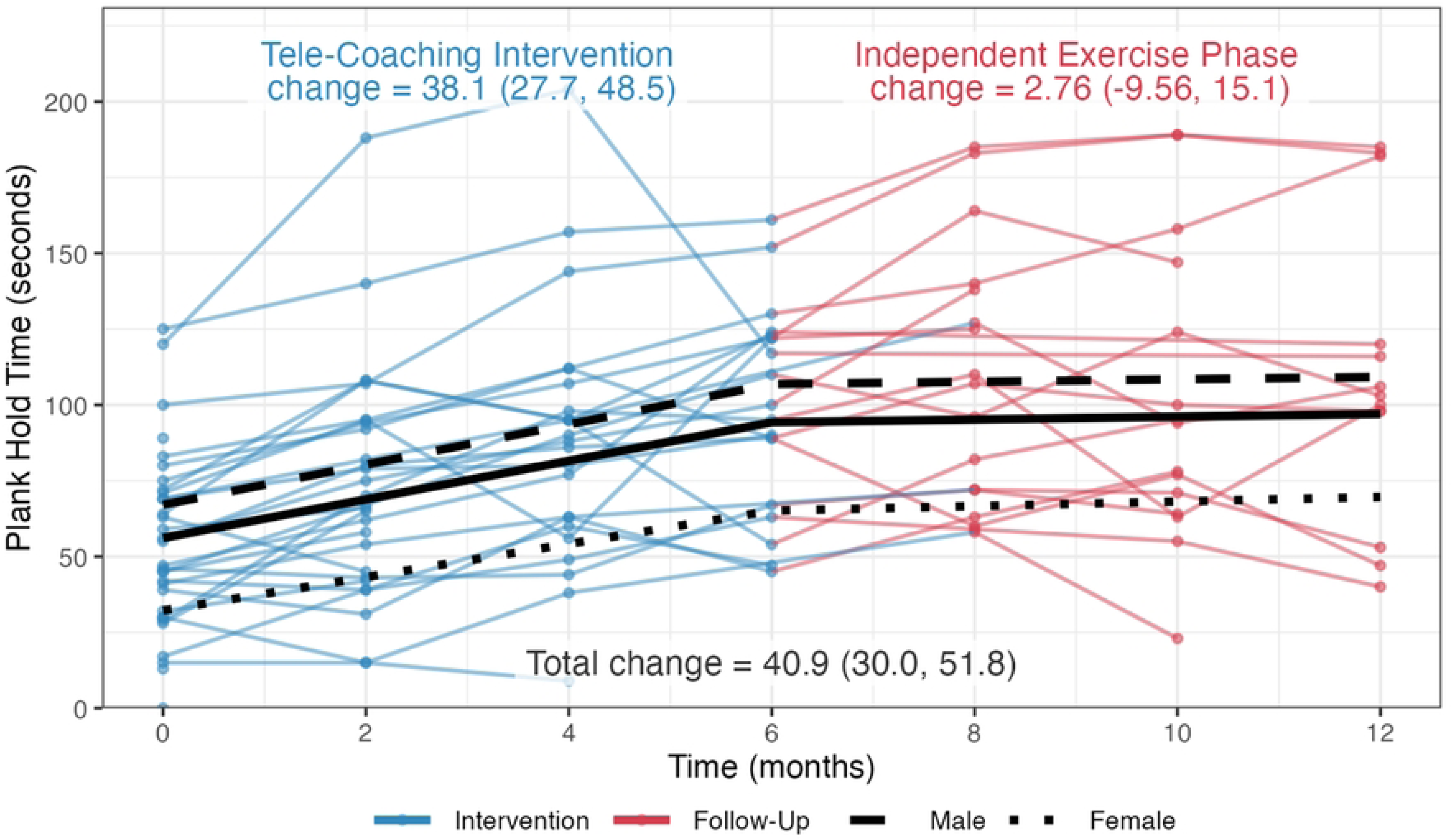
– Strength Outcome – Plank time (seconds)

**Figure 16.**
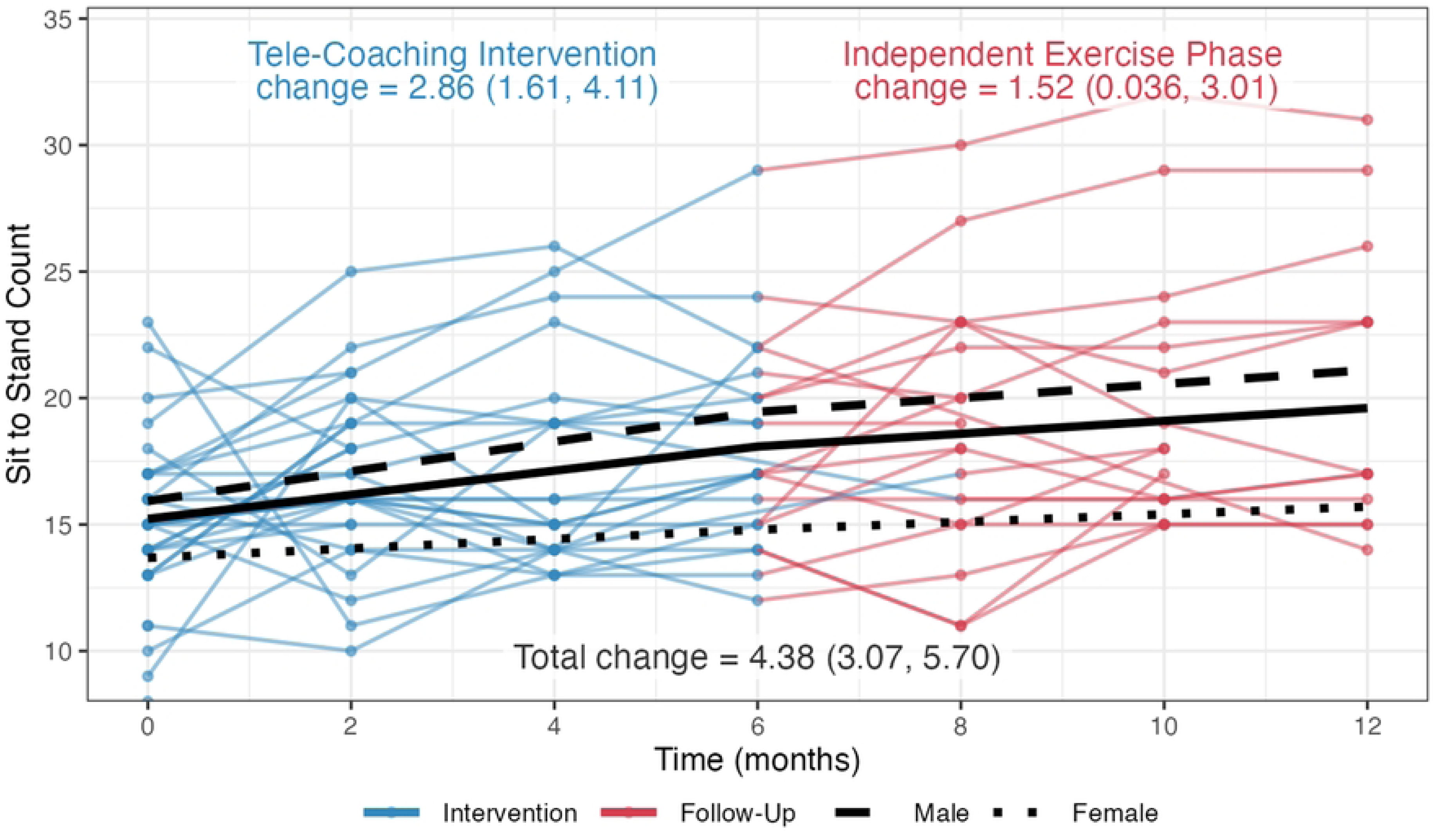
– Physical Function Outcome – Sit-to-stand (number of times in 30 seconds)

**Figure 17.**
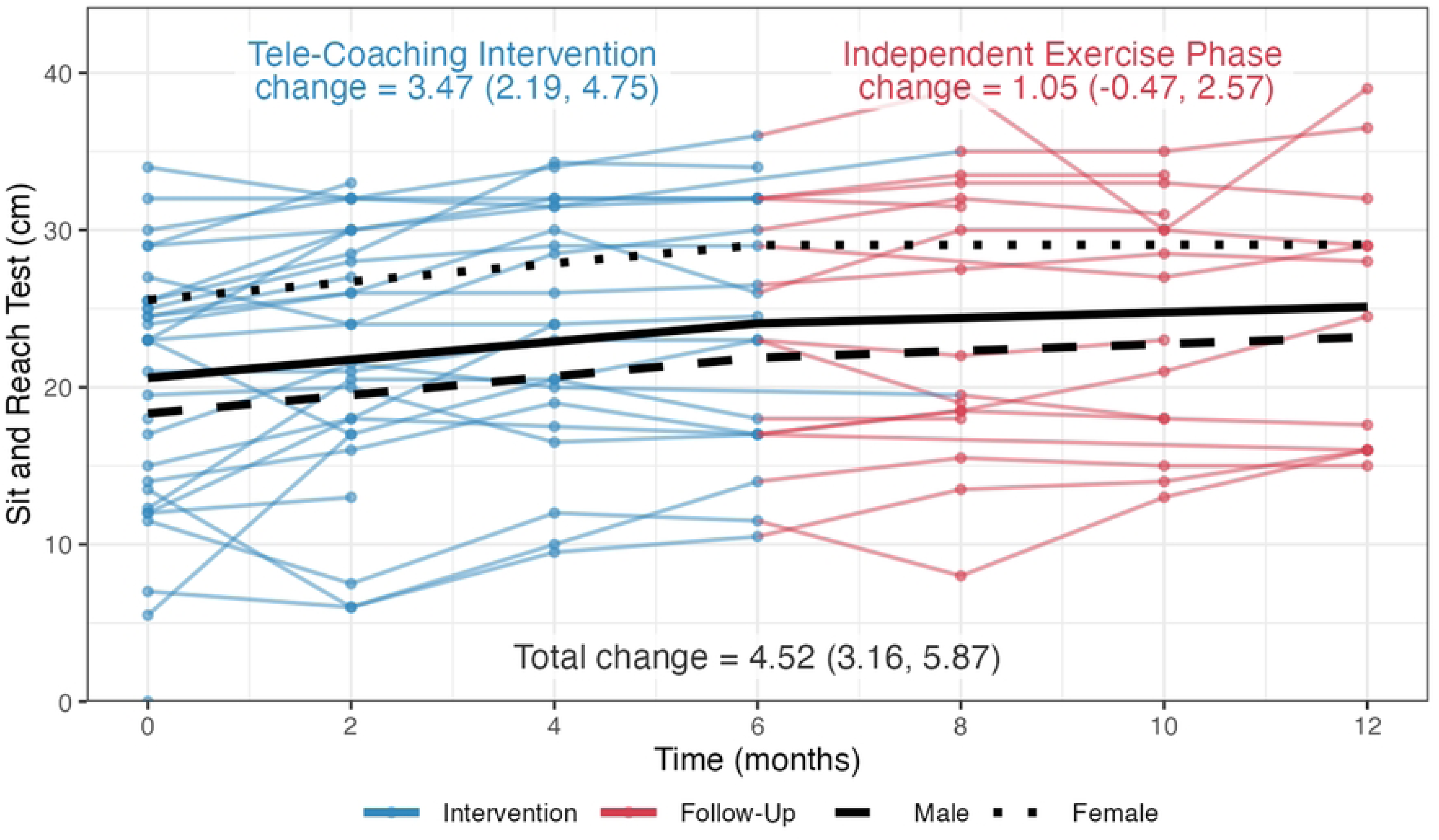
– Flexibility Outcome – Sit-and-reach test (cm)

## Discussion

Results from this study describe the nature of engagement, and effect of an online tele-coaching intervention with adults living with HIV. Results demonstrated increased engagement in physical activity, and improvements in outcomes of cardiopulmonary fitness, weight and body composition, strength, physical function, and flexibility for adults living with HIV in the study.

Characteristics of the participants in this study represented a median age of 53 years, 50 years of age or older (59%), taking antiretroviral therapy with an undetectable viral load (91%). Characteristics of participants in this study are similar to persons living with HIV engaged in care in Ontario [61], with a slight greater representation of females (28%) compared to the estimated 25% of persons living with HIV in Canada in 2022 [62]. While 100 individuals indicated they were interested in learning more about the study, 37 met with the research coordinator to screen for eligibility, of which 32 enrolled and initiated the intervention, which may reflect our recruitment occurring during the course of the COVID-19 pandemic (May to November 2021) and the level of commitment required with the 12-month two-phased exercise intervention and frequency of assessments. The overall retention rate in this study (69% at end of intervention; 56% at study completion) was lower than reported in an earlier in-person CBE intervention study at the YMCA, whereby 84% of participants (67/80) completed a six-month intervention, of which 77% (52/67) completed an 8-month follow-up phase [11]. The in-person CBE intervention study was conducted prior to the COVID-19 pandemic and involved in-person questionnaire administration and fitness assessments, which may have influenced the level of retention in the study [11]. While in-person exercise interventions tend to report greater adherence than online exercise, online interventions provide promise for offering flexibility for enhancing engagement in physical activity for adults living with HIV who may experience episodic disability and face barriers accessing traditional exercise environments [63–65].

### Engagement in Physical Activity

There was an increase in engagement in weekly physical activity and achievement of aerobic physical activity guidelines during the intervention, that were subsequently diminished in the follow-up phase. There was variability in days engaged in physical activity in the past week within and between participants throughout the study (**Fig 2**). Our intention with the combination of group and individualized exercise was to tailor the intervention to individual preferences, while promoting benefits of efficacy for exercise and social support that come from group activity [66]. While engagement in the one-on-one personal training sessions met our threshold for adherence of engaging in ≥75% of the three weekly exercise sessions throughout [55], online group-based exercise classes were less utilized by participants. Interviews with a sub-sample of participants in this study (n=13) highlighted the importance of one-on-one supervision and support from qualified personal trainers as critical to their engagement in the intervention and physical activity [32]. Earlier qualitative inquiry also highlighted one-on-one personal training and supervision a key component to maximizing engagement in physical activity from persons living with HIV and fitness trainers [12, 13]. Our intervention included features with the aim to reduce barriers to engaging in exercise such as providing an online gym membership (providing access to the online group classes), personalized fitness instruction, and monthly self-management educational sessions; we also allowed participants to reschedule their weekly coaching sessions to accommodate for episodes of illness [31]. Nevertheless, despite continued access to the online YMCA membership, providing access to online group exercise classes, the benefits of engagement in days of physical activity and achievement in aerobic and resistance guidelines were reduced during the independent follow-up phase highlighting the importance of one-on-one supervision to sustain engagement in physical activity. Reduced physical activity in the follow-up independent phase of this study was similarly found in our earlier work involving weekly one-on-one supervision with fitness trainers in-person at the YMCA during the intervention, with the expectation that persons would continue to exercise thrice weekly independently in the follow up phase [11]. This may suggest that supportive supervision once every two weeks during an intervention phase can result in similar outcomes of sustained physical activity with less resources than required with weekly supervision, suggesting promise for future scalability and feasibility.

Examining the role of peer-support and alternative self-management approaches as less costly features of CBE interventions are important to promote sustained uptake and benefits on health outcomes for persons aging with HIV [67–69]. Future research also may explore the ideal timeframe needed to foster changes to sustain independent exercise, influence of environmental, personal, and social factors on engagement with online CBE and identify personalized tailored strategies to optimize sustained engagement in CBE interventions for persons living with HIV.

### Strength, weight, body composition, physical function, and flexibility

We observed significant improvements for some outcomes over the six-month intervention, and during the independent follow-up phase, resulting in sustained improvements for weight and body composition, strength, physical function, and flexibility outcomes. Improvements in the 30 second sit to stand test of +4.38 times (improvement from a mean of 15.22 times at baseline to 19.60 times at study completion) surpassed a minimum clinical importance difference (MCID) established in persons with chronic obstructive pulmonary disease (2 or more repetitions) suggesting a potential clinically important improvement of strength, endurance, and physical function [70]. Total number of sit-to-stands in 30 seconds among this sample of participants at baseline were similar to those reported in community-dwelling older adults, 60-69 years (mean of 14 times; +/-2.4) years) [71]. Improvements in strength as measured by the increase in number of push-ups completed in 1 minute (+8.61 push-ups) and flexibility as measured by the sit and reach test (+4.52 cm) surpassed improvements reported in our in-person CBE study (+2.30 push-ups; +1.74 cm on sit-and-reach test)[11], and were similar to improvements in flexibility (+4.1 cm; sit and reach test) after a physiotherapy-led group rehabilitation intervention with adults living with HIV in the United Kingdom [72]. Improvements in BMI and waist circumference found among participants in this study are important as these outcomes in combination are associated with reducing morbidity and mortality among men and women [53], and of particular importance for persons living with HIV who experience weight gain with antiretroviral therapy, and may be at risk of metabolic syndrome, type 2 diabetes, and cardiovascular disease [73–76]. Despite the significant reduction and undesirable outcomes of mean fat free mass (-0.80 kg) and muscle mass (-0.65 kg) at study completion, the rate of change (slope) in these outcomes was not significant across both phases of the study (**Supplemental File 5).** Lower muscle mass is associated with functional impairment among adults aging with HIV [77–79]. Muscle strength and physical performance is an important part of screening for and managing sarcopenia in adults living with HIV [80]. Exercise, particularly resistance training has a role for increasing strength, muscle mass and physical function which can reduce the risk of sarcopenia for persons aging with HIV [80–82]. With the prevalence of frailty occurring earlier in people aging with HIV, it is critical to consider the increased risk of sarcopenia and frailty among persons living with HIV [83]. As more persons with HIV live longer, it is important for clinicians to comprehensively assess for frailty, and to consider tailored interventions to enhance strength, muscle mass, and physical function to reduce the risk of frailty among persons aging with HIV [84–87].

### Strengths and Limitations

Strengths of this study included our evaluation of implementation of an online CBE intervention for adults living with HIV using an Implementation Science approach with the RE-AIM Framework [30]. This study involved a community-engaged approach with people living with HIV, representatives from community-based organizations, clinicians working in HIV care, researchers and recreation fitness providers.

Benefits to physical activity, fitness and health outcomes were attributed to participants who remained in the study. Variability within and between participants for some outcomes, may reflect the fluctuating and potentially episodic nature of HIV as well as the potential for measurement error given the fitness assessments were conducted online via Zoom [88]. Clinical importance of outcomes, particularly related to weight and body composition outcomes are less clear as changes in outcomes will be dependent on baseline measures and sex of participants (women had lower fat free mass and muscle mass) (**Figure 9 and 10**). Measures of physical activity were self-report which may have resulted in measurement error; and only 56% of participants used the WPAM. Report of physical activity among this sub-group of participants is beyond the scope of this paper and an area of future inquiry. We were not powered to detect differences in outcomes by sex; nevertheless, we provide results from our exploratory analyses by sex for future consideration with online CBE (**Supplemental File 6 and 7**).

Challenges implementing the online CBE intervention included recruitment and retention of participants across the 12-month long study during the COVID-19 pandemic. Given the pilot nature of the study design, our analysis is descriptive and exploratory in nature. Findings will inform larger scale implementation of online CBE interventions for people living with HIV. Lastly, this study was conducted in an urban community fitness centre in Toronto, Canada. Participants who enrolled in the study were mostly linked to HIV care, were engaged in some form of exercise at baseline (59%), and may have been more likely to possess a higher level of awareness of self-health compared to the broader HIV population. Generalizability of findings to the broader HIV population, other fitness centres, and in other contexts such as low-and middle-income countries is unclear.

## Conclusions

Adults living with HIV who remained engaged in the 12-month two-phased online community-based exercise intervention study demonstrated increases in physical activity and improvements in strength, weight, body composition, physical function and flexibility outcomes during the online intervention. Future CBE interventions should incorporate strategies to enhance access to, and engagement in physical activity across persons living with episodic illness such as HIV.

## Data Availability

Data are not openly available as participants did not consent to providing public access to their data in the consent process. All relevant data supporting the findings are within the manuscript and its Supporting Information files.

## Acknowledgments

We thank the participants for their contributions to this study and the community organizations who were engaged in this research. Thank you to Raj Maharaj, Maple Leaf Medical Clinic for their support with recruitment via the Ontario HIV Treatment Network Cohort Study (OCS) for this study. We acknowledge the following who were involved in acquisition of funding, protocol development and data collection: Tizneem Jiancaro, Brittany Torres, Shaz Islam, Joanne Lindsay, and Colleen Price.

## Supporting Information

**S1 File. Supplemental File 1 –** Statistical Supplement

**S2 File. Supplemental File 2 -** Characteristics of Participants Who Completed and Did not Complete the Study

**S3 File. Supplemental File 3** – Access to Technology and Space for the Online CBE Intervention

**S4 File. Supplemental File 4** - Engagement in Components of the Online Community-Based Exercise Intervention

**S5 File. Supplemental File 5** - Rate of Change (Slope) in Outcomes during the Intervention and Follow-Up Independent Phases of the Tele-coaching CBE Intervention

**S6 File. Supplemental File 6 -** Exploratory Sex Analyses – Outcomes at each phase of study and Change in Outcomes by Sex

**S7 File. Supplemental File 7** - Sex Analyses (Exploratory) - Rate of Change (Slope) in the Outcomes by Sex

## References

1. Deeks SG, Lewin SR, Havlir DV. The end of AIDS: HIV infection as a chronic disease. Lancet. 2013;382(9903):1525–33. Epub 2013/10/25. doi: 10.1016/S0140-6736(13)61809-7. PubMed PMID: 24152939.

2. Guaraldi G, Prakash M, Moecklinghoff C, Stellbrink HJ. Morbidity in older HIV-infected patients: impact of long-term antiretroviral use. AIDS Rev. 2014;16(2):75–89. PubMed PMID: 24759453.

3. Kendall CE, Wong J, Taljaard M, Glazier RH, Hogg W, Younger J, et al. A cross-sectional, population-based study measuring comorbidity among people living with HIV in Ontario. BMC public health. 2014;14(1):161. Epub 2014/02/15. doi: 10.1186/1471-2458-14-161. PubMed PMID: 24524286; PubMed Central PMCID: PMC3933292.

4. Caspersen CJ, Powell KE, Christenson GM. Physical activity, exercise, and physical fitness: definitions and distinctions for health-related research. Public Health Rep. 1985;100(2):126–31. PubMed PMID: 3920711; PubMed Central PMCID: PMCPMC1424733.

5. O’Brien KK, Tynan AM, Nixon SA, Glazier RH. Effectiveness of Progressive Resistive Exercise (PRE) in the context of HIV: systematic review and meta-analysis using the Cochrane Collaboration protocol. BMC infectious diseases. 2017;17(1):268. doi: 10.1186/s12879-017-2342-8. PubMed PMID: 28403830; PubMed Central PMCID: PMCPMC5389006.

6. O’Brien KK, Tynan AM, Nixon SA, Glazier RH. Effectiveness of aerobic exercise for adults living with HIV: systematic review and meta-analysis using the Cochrane Collaboration protocol. BMC infectious diseases. 2016;16(1):182. doi: 10.1186/s12879-016-1478-2. PubMed PMID: 27112335; PubMed Central PMCID: PMCPMC4845358.

7. Vancampfort D, Mugisha J, De Hert M, Probst M, Firth J, Gorczynski P, et al. Global physical activity levels among people living with HIV: a systematic review and meta-analysis. Disabil Rehabil. 2018;40(4):388–97. Epub 2016/12/09. doi: 10.1080/09638288.2016.1260645. PubMed PMID: 27929355.

8. Sousa CV, Lewis JE, Simoes HG, Campbell CSG, Zaldivar S, Rodriguez A, et al. The effectiveness of a community-based exercise program on depression symptoms among people living with HIV. AIDS Care. 2020:1–7. Epub 2020/01/31. doi: 10.1080/09540121.2020.1719278. PubMed PMID: 31996018.

9. Salbach NM, Howe JA, Brunton K, Salisbury K, Bodiam L. Partnering to Increase Access to Community Exercise Programs for People With Stroke, Acquired Brain Injury and Multiple Sclerosis. Journal of physical activity & health. 2013. Epub 2013/05/17. PubMed PMID: 23676952.

10. Lee LL, Arthur A, Avis M. Using self-efficacy theory to develop interventions that help older people overcome psychological barriers to physical activity: a discussion paper. Int J Nurs Stud. 2008;45(11):1690–9. Epub 2008/05/27. doi: 10.1016/j.ijnurstu.2008.02.012. PubMed PMID: 18501359.

11. O’Brien KK, Davis AM, Chan Carusone S, Avery L, Tang A, Solomon P, et al. Examining the impact of a community-based exercise intervention on cardiorespiratory fitness, cardiovascular health, strength, flexibility and physical activity among adults living with HIV: A three-phased intervention study. PloS one. 2021;16(9):e0257639. Epub 2021/09/25. doi: 10.1371/journal.pone.0257639. PubMed PMID: 34559851; PubMed Central PMCID: PMCPMC8462727.

12. Solomon P, Chan Carusone S, Davis AM, Aubry R, O’Brien KK. A Qualitative Study of Fitness Coaches’ Experiences in Community Based Exercise with People Living with HIV. Journal of the International Association of Providers of AIDS Care. 2021;20:23259582211046762. Epub 2021/10/21. doi: 10.1177/23259582211046762. PubMed PMID: 34668422; PubMed Central PMCID: PMCPMC8532256.

13. Solomon P, Carusone SC, Davis AM, Aubry R, O’Brien KK. Experiences of People Living With HIV in Community Based Exercise: A Qualitative Longitudinal Study. Journal of the International Association of Providers of AIDS Care. 2021;20:2325958221995344. Epub 2021/02/23. doi: 10.1177/2325958221995344. PubMed PMID: 33611978; PubMed Central PMCID: PMC7903824.

14. O’Brien KK, Bayoumi AM, Solomon P, Tang A, Murzin K, Chan Carusone S, et al. Evaluating a community-based exercise intervention with adults living with HIV: protocol for an interrupted time series study. BMJ Open. 2016;6(10):e013618. Epub 2016/11/01. doi: 10.1136/bmjopen-2016-013618. PubMed PMID: 27798038; PubMed Central PMCID: PMC5073553.

15. Bettger JP, Thoumi A, Marquevich V, Groote WD, Battistella LR, Imamura M, et al. COVID-19: maintaining essential rehabilitation services across the continuum. BMJ Global Health 2020;5.

16. Winward C. Supporting community-based exercise in long-term neurological conditions: experience from the Long-term Individual Fitness Enablement (LIFE) project. Clin Rehabil. 2011;25(7):579–87. Epub 2011/06/24. doi: 10.1177/0269215510392075. PubMed PMID: 21697208.

17. Pang MY, Eng JJ, Dawson AS, McKay HA, Harris JE. A community-based fitness and mobility exercise program for older adults with chronic stroke: a randomized, controlled trial. J Am Geriatr Soc. 2005;53(10):1667–74. Epub 2005/09/27. doi: 10.1111/j.1532-5415.2005.53521.x. PubMed PMID: 16181164; PubMed Central PMCID: PMC3226792.

18. D’Hooghe M, Van Gassen G, Kos D, Bouquiaux O, Cambron M, Decoo D, et al. Improving fatigue in multiple sclerosis by smartphone-supported energy management: The MS TeleCoach feasibility study. Multiple sclerosis and related disorders. 2018;22:90–6. Epub 2018/04/13. doi: 10.1016/j.msard.2018.03.020. PubMed PMID: 29649789.

19. Booth G, Zala S, Mitchell C, Zarnegar R, Lucas A, Gilbert AW. The patient acceptability of a remotely delivered pain management programme for people with persistent musculoskeletal pain: A qualitative evaluation. British Journal of Pain. 0(0):20494637221106411. doi: 10.1177/20494637221106411.

20. Loeckx M, Rabinovich RA, Demeyer H, Louvaris Z, Tanner R, Rubio N, et al. Smartphone-Based Physical Activity Telecoaching in Chronic Obstructive Pulmonary Disease: Mixed-Methods Study on Patient Experiences and Lessons for Implementation. JMIR Mhealth Uhealth. 2018;6(12):e200. Epub 2018/12/24. doi: 10.2196/mhealth.9774. PubMed PMID: 30578215; PubMed Central PMCID: PMCPMC6320438.

21. Demeyer H, Louvaris Z, Frei A, De Jong C, Loeckx M, Buesching G, et al. Tele-coaching to promote physical activity in patients with COPD: Evaluation by patients. European Respiratory Journal. 2015;46(Suppl 59):PA3292.

22. Effing T, Zielhuis G, Kerstjens H, van der Valk P, van der Palen J. Community based physiotherapeutic exercise in COPD self-management: a randomised controlled trial. Respiratory medicine. 2011;105(3):418–26. Epub 2010/10/19. doi: 10.1016/j.rmed.2010.09.017. PubMed PMID: 20951018.

23. Castle EM, Dijk G, Asgari E, Shah S, Phillips R, Greenwood J, et al. The Feasibility and User-Experience of a Digital Health Intervention Designed to Prevent Weight Gain in New Kidney Transplant Recipients - The ExeRTiOn2 Trial. Frontiers in Nutrition. 2022. doi: 10.3389/fnut.2022.887580.

24. Greenwood SA, Young HML, Briggs J, Castle EM, Walklin C, Haggis L, et al. Evaluating the effect of a digital health intervention to enhance physical activity in people with chronic kidney disease (Kidney BEAM): a multicentre, randomised controlled trial in the UK. Lancet Digit Health. 2024;6(1):e23–e32. Epub 20231114. doi: 10.1016/S2589-7500(23)00204-2. PubMed PMID: 37968170.

25. Walklin CG, Young HML, Asghari E, Bhandari S, Billany RE, Bishop N, et al. The effect of a novel, digital physical activity and emotional well-being intervention on health-related quality of life in people with chronic kidney disease: trial design and baseline data from a multicentre prospective, wait-list randomised controlled trial (kidney BEAM). BMC nephrology. 2023;24(1):122. Epub 20230502. doi: 10.1186/s12882-023-03173-7. PubMed PMID: 37131125; PubMed Central PMCID: PMCPMC10152439.

26. Young HML, Castle EM, Briggs J, Walklin C, Billany RE, Asgari E, et al. The development and internal pilot trial of a digital physical activity and emotional well-being intervention (Kidney BEAM) for people with chronic kidney disease. Scientific reports. 2024;14(1):700. Epub 20240106. doi: 10.1038/s41598-023-50507-4. PubMed PMID: 38184737; PubMed Central PMCID: PMCPMC10771473.

27. Greenwood SA, Briggs J, Walklin C, Mangahis E, Young HML, Castle EM, et al. Kidney Beam-A Cost-Effective Digital Intervention to Improve Mental Health. Kidney Int Rep. 2024;9(11):3204–17. Epub 20240902. doi: 10.1016/j.ekir.2024.08.030. PubMed PMID: 39534205; PubMed Central PMCID: PMCPMC11551101.

28. Alley S, Jennings C, Plotnikoff RC, Vandelanotte C. Web-Based Video-Coaching to Assist an Automated Computer-Tailored Physical Activity Intervention for Inactive Adults: A Randomized Controlled Trial. J Med Internet Res. 2016;18(8):e223. Epub 2016/08/16. doi: 10.2196/jmir.5664. PubMed PMID: 27520283; PubMed Central PMCID: PMC5002066.

29. Lambert TE, Harvey LA, Avdalis C, Chen LW, Jeyalingam S, Pratt CA, et al. An app with remote support achieves better adherence to home exercise programs than paper handouts in people with musculoskeletal conditions: a randomised trial. Journal of physiotherapy. 2017;63(3):161–7. Epub 2017/07/01. doi: 10.1016/j.jphys.2017.05.015. PubMed PMID: 28662834.

30. Glasgow RE, Eckstein ET, Elzarrad MK. Implementation science perspectives and opportunities for HIV/AIDS research: integrating science, practice, and policy. J Acquir Immune Defic Syndr. 2013;63 Suppl 1:S26–31. Epub 2013/05/17. doi: 10.1097/QAI.0b013e3182920286. PubMed PMID: 23673882.

31. O’Brien KK, Ibanez-Carrasco F, Carusone SC, Bayoumi AM, Tang A, McDuff K, et al. Piloting an online telecoaching community-based exercise intervention with adults living with HIV: protocol for a mixed-methods implementation science study. BMJ Open. 2023;13(3):e067703. Epub 20230330. doi: 10.1136/bmjopen-2022-067703. PubMed PMID: 36997255; PubMed Central PMCID: PMCPMC10069544.

32. Ibanez-Carrasco F, McDuff, K., Da Silva, G., Bayoumi, A.M., Chan Carusone, S., Loutfy, M., Tang, A., O’Brien, K.K. Qualitative insights from an online community-based exercise intervention for persons living with HIV. Front Rehabil Sci, Disability, Rehabilitation, and Inclusion. 2025;6. doi: 10.3389/fresc.2025.1602007.

33. Su TT, Chan Carusone, S., McDuff, K., Ibanez-Carrasco, F., Tang, A., Bayoumi, A.M., Loutfy, M., Avery, L., Da Silva, G., Furlan, A., Trent, H., Ilic, I., Pandovski, Z., Zobeiry, M., Ahluwalia, P., Krizmancic, K., Jiancaro, T., Torres, B., Solomon, P., O’Brien, K.K. Goals in Motion: Exploring goal setting among adults living with HIV who participated in an online community-based exercise intervention. Frontiers in Rehabilitation Sciencdes. 2025;6:1644139. doi: doi: 10.3389/fresc.2025.1644139.

34. Montgomery CA, Henning KJ, Kantarzhi SR, Kideckel TB, Yang CF, O’Brien KK. Experiences participating in a community-based exercise programme from the perspective of people living with HIV: a qualitative study. BMJ Open. 2017;7(4):e015861. Epub 2017/04/06. doi: 10.1136/bmjopen-2017-015861. PubMed PMID: 28377397; PubMed Central PMCID: PMC5387963.

35. Li A, McCabe T, Silverstein E, Dragan S, Salbach NM, Zobeiry M, et al. Community-Based Exercise in the Context of HIV. Journal of the International Association of Providers of AIDS Care. 2017:2325957416686836. doi: 10.1177/2325957416686836. PubMed PMID: 28074681.

36. Zoom Health Care Video Communications Inc. Zoom Health Care Video Communications. 2026.

37. Warburton DER, Jamnik VK, Bredin SSD, Gledhill N, on behalf of the PAR-Q+ Collaboration. The Physical Activity Readiness Questionnaire for Everyone (PAR-Q+) and Electronic Physical Activity Readiness Medical Examination (ePARmed-X+). Health & Fitness Journal of Canada. 2011;4(2):3–23.

38. Rourke SB, Gardner S, Burchell AN, Raboud J, Rueda S, Bayoumi AM, et al. Cohort profile: the Ontario HIV Treatment Network Cohort Study (OCS). International journal of epidemiology. 2013;42(2):402–11. doi: 10.1093/ije/dyr230. PubMed PMID: 22345312.

39. Surveillance and Epidemiology Division, Professional Guidelines and Public Health Practice Division, Centre for Communicable Disease and Infection Control, Canada PHAo. Summary: Estimates of HIV incidence, prevalence and Canada’s progress on meeting the 90-90-90 HIV targets, 20162021. Available from: https://www.canada.ca/en/public-health/services/publications/diseases-conditions/summary-estimates-hiv-incidence-prevalence-canadas-progress-90-90-90.html.

40. Realize. E-module for evidence-informed HIV rehabilitation. Toronto, Canada. 2026. Available from: http://emodule.realizecanada.org/.

41. Fitbit Inc. Fitbit® Official Site 2026. Available from: http://www.fitbit.com/ca/zip.

42. Dagenais M, Cheng D, Salbach NM, Brooks D, O’Brien KK. Wireless physical activity monitor use among adults living with HIV: a scoping review. Rehabilitation Oncology. 2018;37(1):17–28.

43. Huggins AC, Ritzhaupt AD, Dawson KM. Measuring information and communication technology literacy using a performance assessment: validation of the student tool for technology literacy (ST2L). Computers & Education. 2014;77:1–12. doi: 10.1016/j.compedu.2014.04.00512.

44. International Telecommunication Union. Manual for measuring ICT access and use by households and individuals. Place des Nations, Geneva Switzerland: 2014.

45. Information Telecommunication Union. The ICT Development Index (IDI): Conceptual framework and methodology2011. Available from: https://www.itu.int/en/ITU-D/Statistics/Pages/publications/mis/methodology.aspx.

46. Milton K, Bull FC, Bauman A. Reliability and validity testing of a single-item physical activity measure. British journal of sports medicine. 2011;45(3):203–8. Epub 2010/05/21. doi: 10.1136/bjsm.2009.068395. PubMed PMID: 20484314.

47. Canadian Society for Exercise Physiology. Canadian 24-Hour Movement Guidelines for Adults aged 18-64 years: An Integration of Physical Activity, Sedentary Behaviour, and Sleep. 2025.

48. Topolski TD, LoGerfo J, Patrick DL, Williams B, Walwick J, Patrick MB. The Rapid Assessment of Physical Activity (RAPA) among older adults. Prev Chronic Dis. 2006;3(4):A118. PubMed PMID: 16978493; PubMed Central PMCID: PMCPMC1779282.

49. Noguchi KS, O’Brien KK, Aubry RL, Carusone SC, Avery L, Solomon P, et al. Construct Validity and Responsiveness of the Rapid Assessment of Physical Activity in Adults Living With HIV. Archives of Rehabilitation Research and Clinical Translation. 2021;3(4):100164. doi: 10.1016/j.arrct.2021.100164.

50. Jones CJ, Rikli RE, Beam WC. A 30-s Chair-Stand Test as a Measure of Lower Body Strength in Community-Residing Older Adults. Research Quarterly for Exercise and Sport. 1999;70(2):113–9. doi: 10.1080/02701367.1999.10608028.

51. Santo AS, Golding LA. Predicting Maximum Oxygen Uptake from a Modified 3-Minute Step Test. Research Quarterly for Exercise and Sport. 2003;74(1):110–5. doi: 10.1080/02701367.2003.10609070.

52. Kieu NTV, Jung SJ, Shin SW, Jung HW, Jung ES, Won YH, et al. The Validity of the YMCA 3-Minute Step Test for Estimating Maximal Oxygen Uptake in Healthy Korean and Vietnamese Adults. J Lifestyle Med. 2020;10(1):21–9. doi: 10.15280/jlm.2020.10.1.21. PubMed PMID: 32328445; PubMed Central PMCID: PMCPMC7171059.

53. Ross R, Neeland IJ, Yamashita S, Shai I, Seidell J, Magni P, et al. Waist circumference as a vital sign in clinical practice: a Consensus Statement from the IAS and ICCR Working Group on Visceral Obesity. Nat Rev Endocrinol. 2020;16(3):177–89. Epub 20200204. doi: 10.1038/s41574-019-0310-7. PubMed PMID: 32020062; PubMed Central PMCID: PMCPMC7027970.

54. Wells KF, Dillon EK. The Sit and Reach—A Test of Back and Leg Flexibility. Research Quarterly American Association for Health, Physical Education and Recreation. 1952;23(1):115–8. doi: 10.1080/10671188.1952.10761965.

55. Hicks GE, Benvenuti F, Fiaschi V, Lombardi B, Segenni L, Stuart M, et al. Adherence to a community-based exercise program is a strong predictor of improved back pain status in older adults: an observational study. The Clinical journal of pain. 2012;28(3):195–203. Epub 2011/07/14. doi: 10.1097/AJP.0b013e318226c411. PubMed PMID: 21750458; PubMed Central PMCID: PMC3274640.

56. Abeysinghe T, Balasooriya U, Tsui A. Small-sample forecasting regression or ARIMA models?. Journal of quantitative economics: Journal of the Indian Econometric Society. 2003;1:103–13.

57. Ansari F, Gray K, Nathwani D, Phillips G, Ogston S, Ramsay C, et al. Outcomes of an intervention to improve hospital antibiotic prescribing: interrupted time series with segmented regression analysis. The Journal of antimicrobial chemotherapy. 2003;52(5):842–8. Epub 2003/10/18. doi: 10.1093/jac/dkg459. PubMed PMID: 14563900.

58. Pinheiro JC, Bates, D.M. Mixed effects models in s and s-PLUS. New York: Springer; 2000.

59. Bates D, Maechler, M., Bolker, B., Walker, S. Ime4: Linear-mixed effects models using eigen and S4 [Internet]. 2024.

60. R Core Team. R A Language and Environment for Statistical Computing [Internet] Vienna, Austria: R Foundation for Statistical Computing; 2024.

61. Burchell AN, Gardner S, Light L, Ellis BM, Antoniou T, Bacon J, et al. Implementation and Operational Research: Engagement in HIV Care Among Persons Enrolled in a Clinical HIV Cohort in Ontario, Canada, 2001-2011. J Acquir Immune Defic Syndr. 2015;70(1):e10–9. doi: 10.1097/QAI.0000000000000690. PubMed PMID: 26322672; PubMed Central PMCID: PMCPMC4623844.

62. Public Health Agency of Canada. Canada’s Progress Towards Ending the HIV Epidemic 20222024. Available from: https://www.canada.ca/en/public-health/services/publications/diseases-conditions/canada-progress-towards-ending-hiv-epidemic-2022.html.

63. Oginni J, Otinwa G, Gao Z. Physical Impact of Traditional and Virtual Physical Exercise Programs on Health Outcomes among Corporate Employees. Journal of clinical medicine. 2024;13(3). Epub 20240125. doi: 10.3390/jcm13030694. PubMed PMID: 38337388; PubMed Central PMCID: PMCPMC10856341.

64. Suderman K, Skene T, Sellar C, Dolgoy N, Pituskin E, Joy AA, et al. Virtual or In-Person: A Mixed Methods Survey to Determine Exercise Programming Preferences during COVID-19. Curr Oncol. 2022;29(10):6735–48. Epub 20220920. doi: 10.3390/curroncol29100529. PubMed PMID: 36290806; PubMed Central PMCID: PMCPMC9601145.

65. Monton-Martinez R, Ballester-Ferrer JA, Baladzhaeva S, Sempere-Ruiz N, Casanova-Lizon A, Roldan A, et al. Exploring the Impact of Web-Based vs. In-Person Exercise Training on Benefits and Adherence in Substance Use Disorder Interventions: A Pilot Study. Healthcare (Basel). 2024;12(6). Epub 20240319. doi: 10.3390/healthcare12060684. PubMed PMID: 38540648; PubMed Central PMCID: PMCPMC10969899.

66. Glasgow RE, McKay HG, Piette JD, Reynolds KD. The RE-AIM framework for evaluating interventions: what can it tell us about approaches to chronic illness management? Patient Educ Couns. 2001;44(2):119–27. Epub 2001/08/02. PubMed PMID: 11479052.

67. Burton E, Farrier K, Hill KD, Codde J, Airey P, Hill AM. Effectiveness of peers in delivering programs or motivating older people to increase their participation in physical activity: Systematic review and meta-analysis. Journal of sports sciences. 2018;36(6):666–78. Epub 2017/05/24. doi: 10.1080/02640414.2017.1329549. PubMed PMID: 28535358.

68. Boucher LM, Liddy C, Mihan A, Kendall C. Peer-led Self-management Interventions and Adherence to Antiretroviral Therapy Among People Living with HIV: A Systematic Review. AIDS Behav. 2019. Epub 2019/10/11. doi: 10.1007/s10461-019-02690-7. PubMed PMID: 31598801.

69. Genberg BL, Shangani S, Sabatino K, Rachlis B, Wachira J, Braitstein P, et al. Improving Engagement in the HIV Care Cascade: A Systematic Review of Interventions Involving People Living with HIV/AIDS as Peers. AIDS Behav. 2016;20(10):2452–63. Epub 2016/02/04. doi: 10.1007/s10461-016-1307-z. PubMed PMID: 26837630; PubMed Central PMCID: PMC4970970.

70. Zanini A, Crisafulli E, D’Andria M, Gregorini C, Cherubino F, Zampogna E, et al. Minimum Clinically Important Difference in 30-s Sit-to-Stand Test After Pulmonary Rehabilitation in Subjects With COPD. Respir Care. 2019;64(10):1261–9. Epub 20190703. doi: 10.4187/respcare.06694. PubMed PMID: 31270178.

71. Lein DH, Jr., Alotaibi M, Almutairi M, Singh H. Normative Reference Values and Validity for the 30-Second Chair-Stand Test in Healthy Young Adults. Int J Sports Phys Ther. 2022;17(5):907–14. Epub 20220801. doi: 10.26603/001c.36432. PubMed PMID: 35949374; PubMed Central PMCID: PMCPMC9340829.

72. Brown D, Claffey A, Harding R. Evaluation of a physiotherapy-led group rehabilitation intervention for adults living with HIV: referrals, adherence and outcomes. AIDS Care. 2016:1–11. doi: 10.1080/09540121.2016.1191611. PubMed PMID: 27264319.

73. Lam JO, Leyden WA, Alexeeff S, Lea AN, Hechter RC, Hu H, et al. Changes in Body Mass Index Over Time in People With and Without HIV Infection. Open forum infectious diseases. 2024;11(2):ofad611. Epub 20240206. doi: 10.1093/ofid/ofad611. PubMed PMID: 38323078; PubMed Central PMCID: PMCPMC10846771.

74. Kgatla H, Mokoena H, Ndlovu M, Ziqubu K, Sivhiya MS, Kyeyune J, et al. A systematic review assessing body mass index and waist circumference as predictors for the development of type 2 diabetes mellitus among people living with HIV on antiretroviral therapy. BMC infectious diseases. 2025;25(1):1768. Epub 20251229. doi: 10.1186/s12879-025-12192-8. PubMed PMID: 41462126; PubMed Central PMCID: PMCPMC12752373.

75. Duarte Fernandes EA, Teixeira S, Varela PWA, Alves JCM, de Almeida-Neto PF, Gomes C, et al. Associations of anthropometric indices with body adiposity for assessing cardiovascular risk in people living with HIV: a cross-sectional study. PeerJ. 2025;13:e18833. Epub 20251020. doi: 10.7717/peerj.18833. PubMed PMID: 41142315; PubMed Central PMCID: PMCPMC12548633.

76. Taramasso L, Bonfanti P, Ricci E, Maggi P, Orofino G, Squillace N, et al. Metabolic syndrome and body weight in people living with HIV infection: analysis of differences observed in three different cohort studies over a decade. HIV Med. 2022;23(1):70–9. Epub 20210902. doi: 10.1111/hiv.13165. PubMed PMID: 34473897.

77. Erlandson KM, Allshouse AA, Jankowski CM, MaWhinney S, Kohrt WM, Campbell TB. Functional impairment is associated with low bone and muscle mass among persons aging with HIV infection. J Acquir Immune Defic Syndr. 2013;63(2):209–15. doi: 10.1097/QAI.0b013e318289bb7e. PubMed PMID: 23392468; PubMed Central PMCID: PMCPMC3654048.

78. Martins LC, Oliveira ESMP, Dos Santos ACO, da Silveira VM, de Araujo PSR. Prevalence and associated factors related to sarcopenia in people living with HIV/AIDS. BMC infectious diseases. 2024;24(1):933. Epub 20240909. doi: 10.1186/s12879-024-09845-5. PubMed PMID: 39251940; PubMed Central PMCID: PMCPMC11385512.

79. Gehrke B, Farias MLF, Wildemberg LE, Ferraiuoli GI, Ribeiro V, Bosgnoli R, et al. The importance of muscle strength and physical performance as part of the diagnosis and management of sarcopenia in young adults living with human immunodeficiency virus. Arch Endocrinol Metab. 2025;69(5). Epub 20251009. doi: 10.20945/2359-4292-2025-0018. PubMed PMID: 41066379; PubMed Central PMCID: PMCPMC12510331.

80. SeyedAlinaghi S, Mehraeen E, Mirzapour P, Rahimzadeh P, Abbasi Yazdi A, Roozbahani MM, et al. Effectiveness of exercise on sarcopenia in HIV patients: a systematic review of current literature. AIDS Care. 2025;37(3):349–61. Epub 20250119. doi: 10.1080/09540121.2025.2452528. PubMed PMID: 39828981.

81. Oursler KK, Marconi VC, Briggs BC, Sorkin JD, Ryan AS, Team FVP. Telehealth Exercise Intervention in Older Adults With HIV: Protocol of a Multisite Randomized Trial. J Assoc Nurses AIDS Care. 2022;33(2):168–77. doi: 10.1097/JNC.0000000000000235. PubMed PMID: 33481463; PubMed Central PMCID: PMCPMC8289938.

82. Jankowski CM, Konigsberg IR, Wilson MP, Sun J, Brown TT, Julian CG, et al. Skeletal muscle DNA methylation: Effects of exercise and HIV. Aging Cell. 2024;23(1):e14025. Epub 20231103. doi: 10.1111/acel.14025. PubMed PMID: 37920126; PubMed Central PMCID: PMCPMC10776118.

83. Branas F, Jimenez Z, Sanchez-Conde M, Dronda F, Lopez-Bernaldo De Quiros JC, Perez-Elias MJ, et al. Frailty and physical function in older HIV-infected adults. Age and ageing. 2017;46(3):522–6. doi: 10.1093/ageing/afx013. PubMed PMID: 28203694.

84. Clair-Sullivan NS, Bristowe K, Khan I, Maddocks M, Harding R, Bremner S, et al. Implementation of frailty screening for older people living with HIV in Brighton, UK. HIV Med. 2024;25(4):484–90. Epub 20231207. doi: 10.1111/hiv.13598. PubMed PMID: 38062917.

85. St Clair-Sullivan N, Simmons K, Harding R, Levett T, Maddocks M, Roberts J, et al. Frailty and frailty screening: A qualitative study to elicit perspectives of people living with HIV and their healthcare professionals. HIV Med. 2023;24(4):480–90. Epub 20221013. doi: 10.1111/hiv.13419. PubMed PMID: 36229192.

86. St Clair-Sullivan N, Bristowe K, Bremner S, Maddocks M, Harding R, Levett T, et al. Comprehensive geriatric assessment for people living with HIV and frailty: A mixed-methods feasibility randomized controlled trial. HIV Med. 2025. Epub 20251121. doi: 10.1111/hiv.70149. PubMed PMID: 41271592.

87. Kehler DS, Milic J, Guaraldi G, Fulop T, Falutz J. Frailty in older people living with HIV: current status and clinical management. BMC geriatrics. 2022;22(1):919. Epub 20221130. doi: 10.1186/s12877-022-03477-7. PubMed PMID: 36447144; PubMed Central PMCID: PMCPMC9708514.

88. O’Brien KK, Bayoumi AM, Strike C, Young NL, Davis AM. Exploring disability from the perspective of adults living with HIV/AIDS: development of a conceptual framework. Health Qual Life Outcomes. 2008;6:76. Epub 2008/10/07. doi: 1477-7525-6-76 [pii] 10.1186/1477-7525-6-76. PubMed PMID: 18834538.

